# Understanding how Victoria, Australia gained control of its second COVID-19 wave

**DOI:** 10.1101/2021.04.03.21254866

**Authors:** James M Trauer, Michael J Lydeamore, Gregory W Dalton, David Pilcher, Michael T Meehan, Emma S McBryde, Allen C Cheng, Brett Sutton, Romain Ragonnet

## Abstract

Victoria has been Australia’s hardest hit state by the COVID-19 pandemic, but was successful in reversing its second wave of infections through aggressive policy interventions. The clear reversal in the epidemic trajectory combined with information on the timing and geographical scope of policy interventions offers the opportunity to estimate the relative contribution of each change. We developed a compartmental model of the COVID-19 epidemic in Victoria that incorporated age and geographical structure, and calibrated it to data on case notifications, deaths and health service needs according to the administrative divisions of Victoria’s healthcare, termed clusters. We achieved a good fit to epidemiological indicators, at both the state level and for individual clusters, through a combination of time-varying processes that included changes to case detection rates, population mobility, school closures, seasonal forcing, physical distancing and use of face coverings. Estimates of the risk of hospitalisation and death among persons with disease that were needed to achieve this close fit were markedly higher than international estimates, likely reflecting the concentration of the epidemic in groups at particular risk of adverse outcomes, such as residential facilities. Otherwise, most fitted parameters were consistent with the existing literature on COVID-19 epidemiology and outcomes. We estimated a significant effect for each of the calibrated time-varying processes on reducing the risk of transmission per contact, with broad estimates of the reduction in transmission risk attributable to seasonal forcing (27.8%, 95% credible interval [95%CI] 9.26-44.7% for mid-summer compared to mid-winter), but narrower estimates for the individual-level effect of physical distancing of 12.5% (95%CI 5.69-27.9%) and of face coverings of 39.1% (95%CI 31.3-45.8%). That the multi-factorial public health interventions and mobility restrictions led to the dramatic reversal in the epidemic trajectory is supported by our model results, with the mandatory face coverings likely to have been particularly important.

## Introduction

The COVID-19 pandemic has had an unprecedented impact on human health and society,^1,2^ with high-income, urban and temperate areas often the most severely affected.^3–5^ The impacts of the virus are felt through the direct effect of the virus, particularly through its considerable risk of mortality following infection^6,7^ and likely substantial post-infection sequelae,^8^ but also through the extreme lockdown measures often needed to achieve control.^9^

Australia has been relatively successful in controlling COVID-19,^10^ with all jurisdictions of the country achieving good control of the first wave of imported cases through March and April. However, the southern state of Victoria suffered a substantial second wave of locally-transmitted cases, reaching around 600 notifications per day, predominantly in metropolitan Melbourne in winter.

In response to the pandemic, the Victorian Government implemented a number of recommendations and policy changes with the aim of reversing the escalating case numbers that had a severe impact on social and economic activities. Specific changes included stringent restrictions on movement, increased testing rates, school closures and face covering requirements. In metropolitan Melbourne face coverings were mandated from 23rd July and significantly more stringent movement restrictions were implemented from 9th of July (moving to “stage 3”) and from 2nd August (moving from “stage 3” to “stage 4”). Case numbers peaked in the final days of July and first days of August and declined thereafter. Understanding the relative contribution of each of these interventions is complicated by several interventions being implemented within a few weeks, along with policy differences between metropolitan and regional areas. Nevertheless, the clear reversal in the trajectory of the epidemic following the implementation of these policy changes offers the opportunity to explore the contribution of these factors to the epidemic profile. Indeed, the experience of Victoria’s second wave is virtually unique in that the pattern of substantial and escalating daily community cases was reversed following these policy changes, with elimination subsequently achieved in November. We adapted our computational model to create a unified transmission model for the state and infer the contribution of the policy interventions implemented to changing the direction of the epidemic trajectory.

## Methods

We adapted the transmission dynamic model that we had used to produce forecasts of new cases, health system capacity requirements and deaths for the Victorian Department of Health and Human Services (DHHS) at the health service cluster level (henceforward “cluster”). By incorporating geographical structure to represent clusters, we built a unified model of the COVID-19 epidemic in Victoria, and fitted the model to multiple indicators of epidemic burden in order to infer the effectiveness of each component of the response to the epidemic. Full methods are provided in the Supplementary Methods, key features of the model are illustrated in Figure 1 and all code is available at https://github.com/monash-emu/AuTuMN.

**Figure 1.**
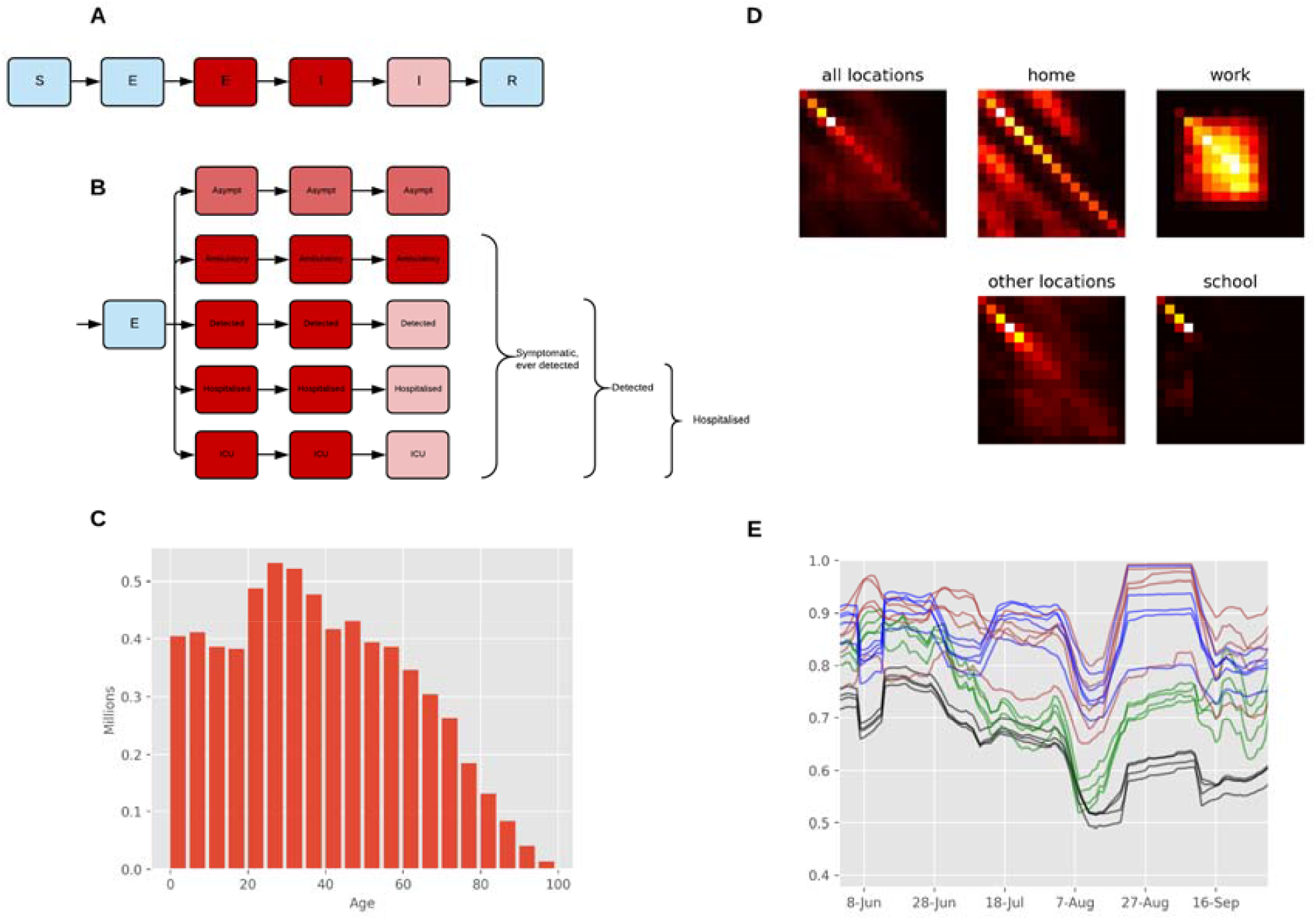
Age-structured COVID-19 model with population distribution, age-specific contact rates, and mobility inputs. (A) Unstratified model structure. (B) Stratification by infection and detection status. (Note that age stratification consists of further stratifying all compartments 16 times.) (C) Starting population age distribution. (D) Heterogeneous mixing matrices by age in the absence of non-pharmaceutical interventions. (E) Macro-distancing adjustments to the mixing matrices for each cluster smoothed with 7-day moving average. Black, workplace mobility for metropolitan clusters; green, other locations mobility for metropolitan clusters; blue, workplace mobility for regional clusters; brown, other locations mobility for regional clusters.

### Base model

Our model of COVID-19 epidemiology is a stratified, deterministic SEIR framework, with sequential compartments representing non-infectious and infectious incubation periods and early and late active disease (Figure 1A). The late incubation compartment and the two active compartments are stratified to simulate epidemiological considerations including asymptomatic cases,^11^ incomplete detection of symptomatic cases, hospitalisation and ICU admission (Figure 1B). All model compartments were then stratified by age, with susceptibility, the clinical fraction, hospitalisation risk and infection fatality rate modified by age group.^6^ We introduced heterogeneous mixing by age using the synthetic mixing matrix for Australia developed by Prem *et al*. 2017 (Figure 1D).^12^

### Simulation of public health interventions

We simulated movement restrictions (including school closures, business closures and working from home) by varying the relative contribution of three of four locations to the overall mixing matrix (Figure 1E) continuously over time. Using Google mobility data (https://www.google.com/covid19/mobility/) weighted to cluster, we scaled the work contribution with workplace mobility and the contribution from other locations (contacts outside of schools, homes, and work) with an average of mobility from the remaining Google mobility locations other than residential (Figure 1E). We simulated school closures by scaling the school contribution according to the proportion of children attending schools on site. We assumed that schools began transitioning to onsite learning from the 26th of May, at which time 400,000 of 1,018,000 students returned to onsite education. The remaining students were considered to return onsite from the 9th of June, before 90% of students moved to remote learning from the 9th of July, which continued until October.

The term “micro-distancing” is used to refer to behavioural changes that reduce the risk of transmission given an interpersonal contact and so are not captured through data on population mobility (e.g. maintaining physical distance and use of face coverings). Micro-distancing was assumed to reduce the risk of both transmission from index cases and the risk of infection of susceptible persons, with the effect of both physical distancing and face coverings applied to all three non-residential locations. Both the coverage and the effectiveness of each intervention were incorporated, with time-varying functions representing the proportion of the population complying with recommendations over time and constant calibration parameters scaling these functions to represent the effectiveness of the intervention. The profiles of compliance with these two recommendations was estimated by fitting to YouGov data, available at https://github.com/YouGov-Data/covid-19-tracker, with hyperbolic tan functions providing a good fit to data (Supplemental Figures 5 and 6). Because face coverings were mandated ten days later in regional Victoria than metropolitan Melbourne, the face coverings compliance function was delayed by this period for regional clusters, while the physical distancing function was identical for all clusters.

We defined the modelled case detection rate as the proportion of all symptomatic cases that were detected (Figure 1B). We related the case detection rate (CDR, Equation 1) to the number of tests performed using an exponential function, under the assumption that a certain per capita daily testing rate is associated with a specific case detection rate, with this relationship varied during calibration (Supplemental Figure 4):

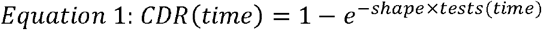

Seasonal forcing was incorporated using a transposed sine function, with maximum value at the winter solstice.

### Incorporation of health service clusters

We further stratified the above model to Victoria’s nine health service clusters, including four clusters which together constitute metropolitan Melbourne (North, West, South, South East Metro) and five regional clusters which together constitute the rest of Victoria (Barwon South West, Gippsland, Grampians, Hume, Loddon-Mallee). We split the estimated age-specific population for Victoria (Figure 1C) according to historical patterns of accessing health service clusters provided by DHHS. The infectious seed was split across the compartments representing current infection and assigned evenly across the metropolitan clusters, with the remainder of the population assigned to the susceptible compartments. The force of infection in each cluster was calculated as a weighted average of the age-specific force of infection for each cluster, with the index cluster having the greatest weight and all non-index clusters having equal weight. The final model included 2,592 compartments interacting through a dynamic mixing matrix of dimensions 144 x 144 (16 age groups and nine geographical patches), with each matrix element scaling over time to reflect changes to population mixing in response to changes in mobility and pandemic-related policy decisions as introduced above.

### Calibration

Because of the high-dimensional parameter space, we calibrated the model to reproduce local COVID-19 dynamics during Victoria’s second wave using an adaptive Metropolis algorithm, which is non-Markovian but retains ergodic properties (Table 1; Supplement).^13^ For the prior distributions of epidemiological calibration parameters, we used uniform priors for highly uncertain quantities and truncated normal distributions for quantities informed by epidemiological evidence. We included adjusters in our calibration parameters to modify the proportion of symptomatic individuals, proportion of symptomatic individuals hospitalised, and the infection fatality rate. The parameters are multiplicative factors that are applied to the odds ratio equivalent to the proportion parameter, rather than directly to the parameter value itself; thus ensuring that the adjusted value lies between zero and one.

The likelihood function was constructed by first incorporating Poisson distributions with rate parameters equal to each of the state-wide daily time-series for notifications, hospitalisations, ICU admissions and deaths. This was then multiplied by terms for the daily time-series of notifications for each cluster, smoothed with a four-day moving average, using normal distributions. As there is no requirement for individuals living in a cluster catchment to attend that health service, we allocated each cluster a proportion of each notification according to the historical tendency of persons from each Local Government Area (LGA) to attend a hospital from that cluster (such that daily cluster-specific notification and death counts are not integer-valued).

**Table 1.** Prior distributions and posterior estimates of all calibrated epidemiological model parameters.

| Parameter (units) | Prior distribution | 2.5th centile | Median | 97.5th centile |
| --- | --- | --- | --- | --- |
| Unadjusted risk of transmission per contact | Uniform, support: [0.015, 0.06] | 0.0254 | 0.0364 | 0.0497 |
| Incubation period (days) | Truncated normal, mean: 5.5, standard deviation: 0.97 support: [1, infinity) | 3.70 | 4.68 | 5.79 |
| Duration of active disease (days) | Truncated normal, mean: 6.5, standard deviation: 0.77 support: [4, infinity) | 4.89 | 5.88 | 7.10 |
| Pre-ICU period (days) | Truncated normal, mean: 12.7, standard deviation: 4 support: [3, infinity) | 5.47 | 11.2 | 17.9 |
| Symptomatic proportion adjuster | Truncated normal, mean: 1, standard deviation: 0.2, support: [0.5, infinity) | 0.715 | 0.984 | 1.35 |
| Infection fatality rate adjuster | Uniform, support: [0.5, 4] | 1.66 | 2.48 | 3.32 |
| Hospitalisation rate adjuster | Uniform, support: [0.5, 3] | 1.44 | 2.31 | 2.90 |
| Infectiousness of asymptomatic persons multiplier | Uniform, support: [0.3, 0.7] | 0.234 | 0.348 | 0.527 |
| Starting infectious population (persons) | Uniform, support: [10, 30] | 25.0 | 40.7 | 61.3 |
| Seasonal forcing | Uniform, support: [0, 0.5] | 0.0926 | 0.278 | 0.447 |
| Case detection rate at one test per 1,000 per day (proportion) | Uniform, support: [0.2, 0.5] | 0.224 | 0.330 | 0.453 |
| Inter-cluster mixing (%) | Uniform, support: [0.005, 0.05] | 0.67 | 1.41 | 2.88 |
| Effect of physical distancing | Uniform, support: [0, 0.5] | 0.0569 | 0.125 | 0.279 |
| Effect of face coverings | Uniform, support: [0, 0.5] | 0.313 | 0.391 | 0.458 |

| Cluster-specific contact rate multipliers |  |  |  |  |
| --- | --- | --- | --- | --- |
| North Metro | Truncated normal,<br>mean: 1,<br>standard deviation: 0.5,<br>support: [0.5, infinity) | 0.768 | 1.24 | 1.83 |
| West Metro |  | 0.924 | 1.46 | 1.99 |
| South Metro |  | 0.659 | 1.07 | 1.55 |
| South East Metro |  | 0.630 | 1.10 | 1.53 |
| Barwon South West |  | 0.635 | 0.988 | 1.47 |
| Other regional clusters |  | 0.533 | 0.679 | 0.996 |

## Results

### Calibration fit

We achieved good calibration fits to all calibration targets (Figures 2-4), along with close matches to cluster-specific indicators not used for calibration (Supplemental Figures 7-10) under the framework of a single state-wide model. The epidemic peaks in the regional clusters occurred somewhat later than in the metropolitan clusters, which is attributable to the modelled infection first being seeded in the metropolitan regions before triggering epidemics outside of Greater Melbourne and is consistent with historical reality. These fits were associated with a post-wave proportion of the population recovered of around 1-2%, with higher proportions in metropolitan regions and young adults (Figure 9).

**Figure 2.**
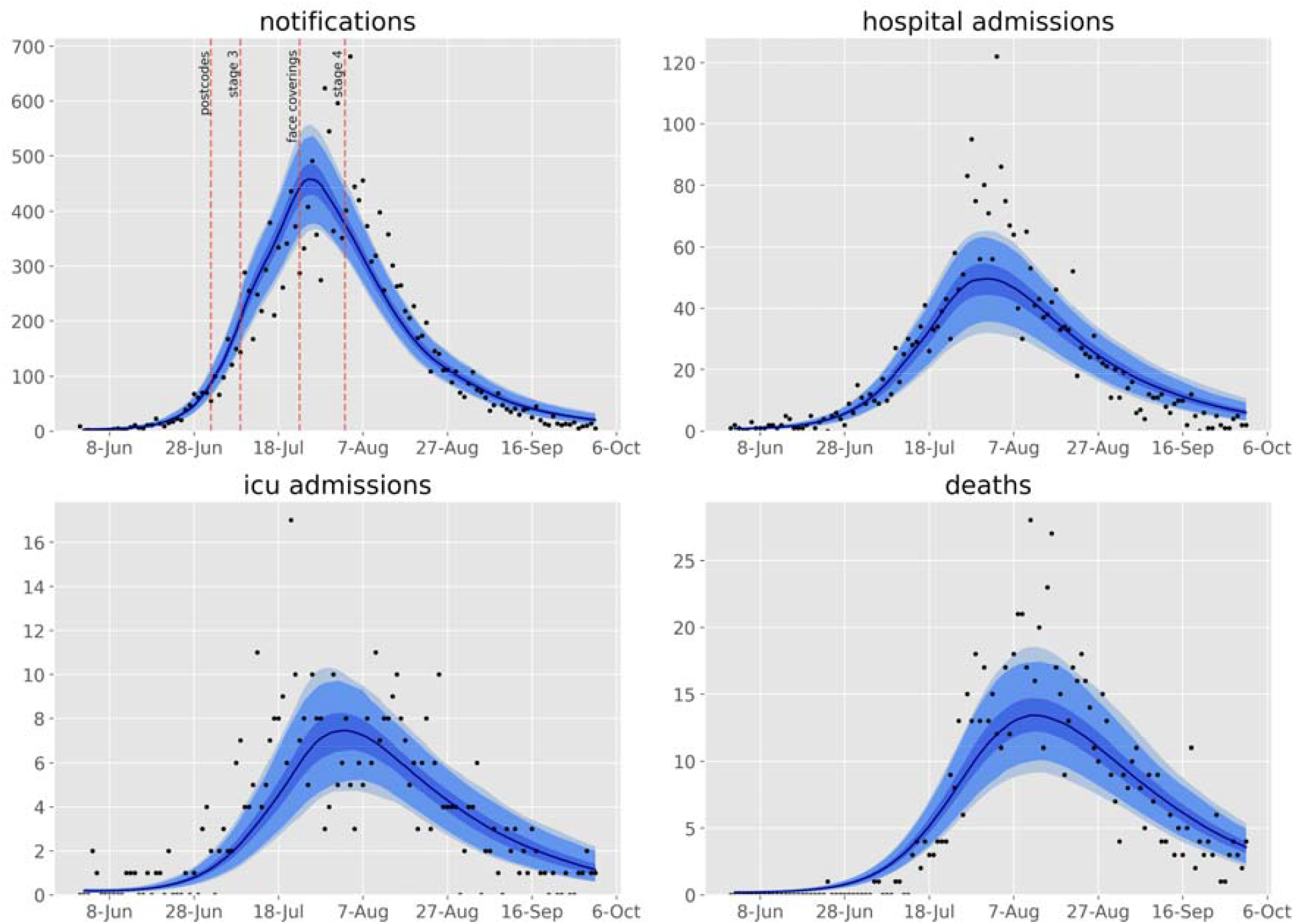
Calibration fits to daily state-wide time series of notifications, hospital admissions, ICU admissions and deaths. Daily confirmed cases (black dots) overlaid on the median modeled detected cases (dark blue line), with shaded areas representing the 25th to 75th centile (mid blue), 2.5th to 97.5th centile (light blue) and 1st to 99th centile (faintest blue) of estimated detected cases. Timing of restrictions applied to metropolitan Melbourne indicated in the upper left panel.

**Figure 3.**
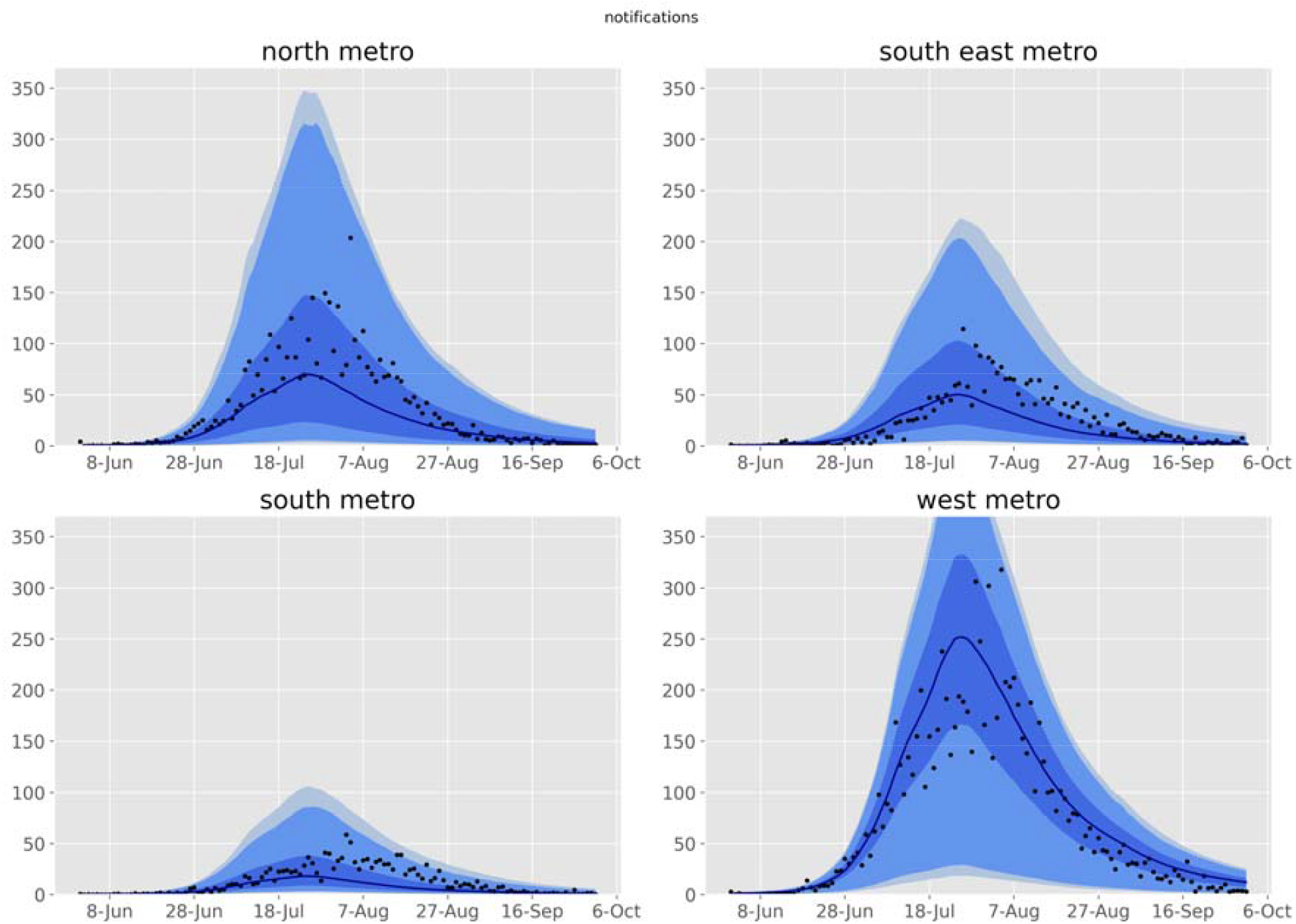
Calibration fits to daily time series of notifications for each metropolitan health service cluster. Daily confirmed cases (black dots) overlaid on the median modeled detected cases (dark blue line), with shaded areas representing the 25th to 75th centile (mid blue), 2.5th to 97.5th centile (light blue) and 1st to 99th centile (faintest blue) of estimated detected cases.

**Figure 4.**
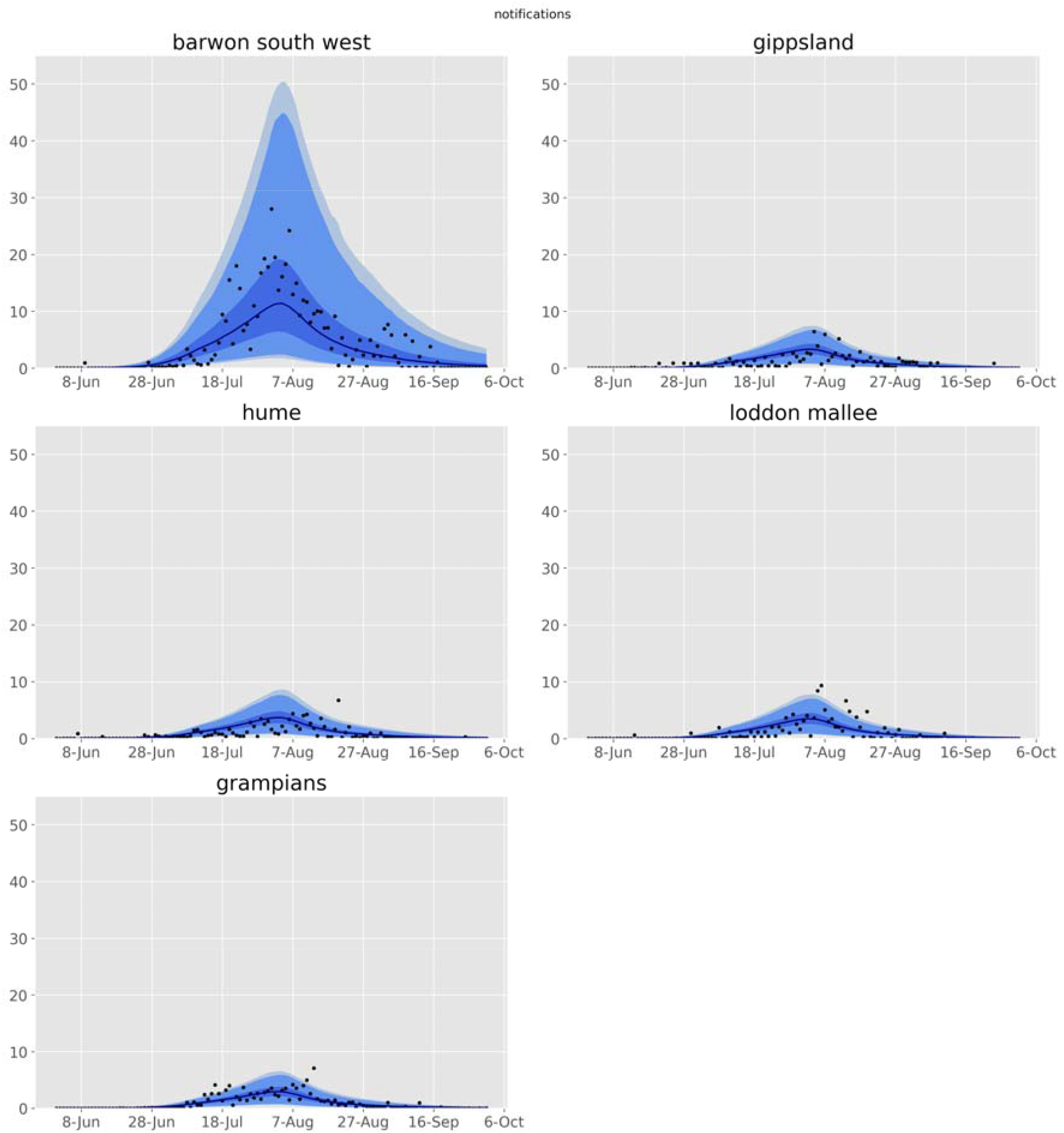
Calibration fits to daily time series of notifications for each regional health service cluster. Daily confirmed cases (black dots) overlaid on the median modeled detected cases (dark blue line), with shaded areas representing the 25th to 75th centile (mid blue), 2.5th to 97.5th centile (light blue) and 1st to 99th centile (faintest blue) of estimated detected cases.

**Figure 5.**
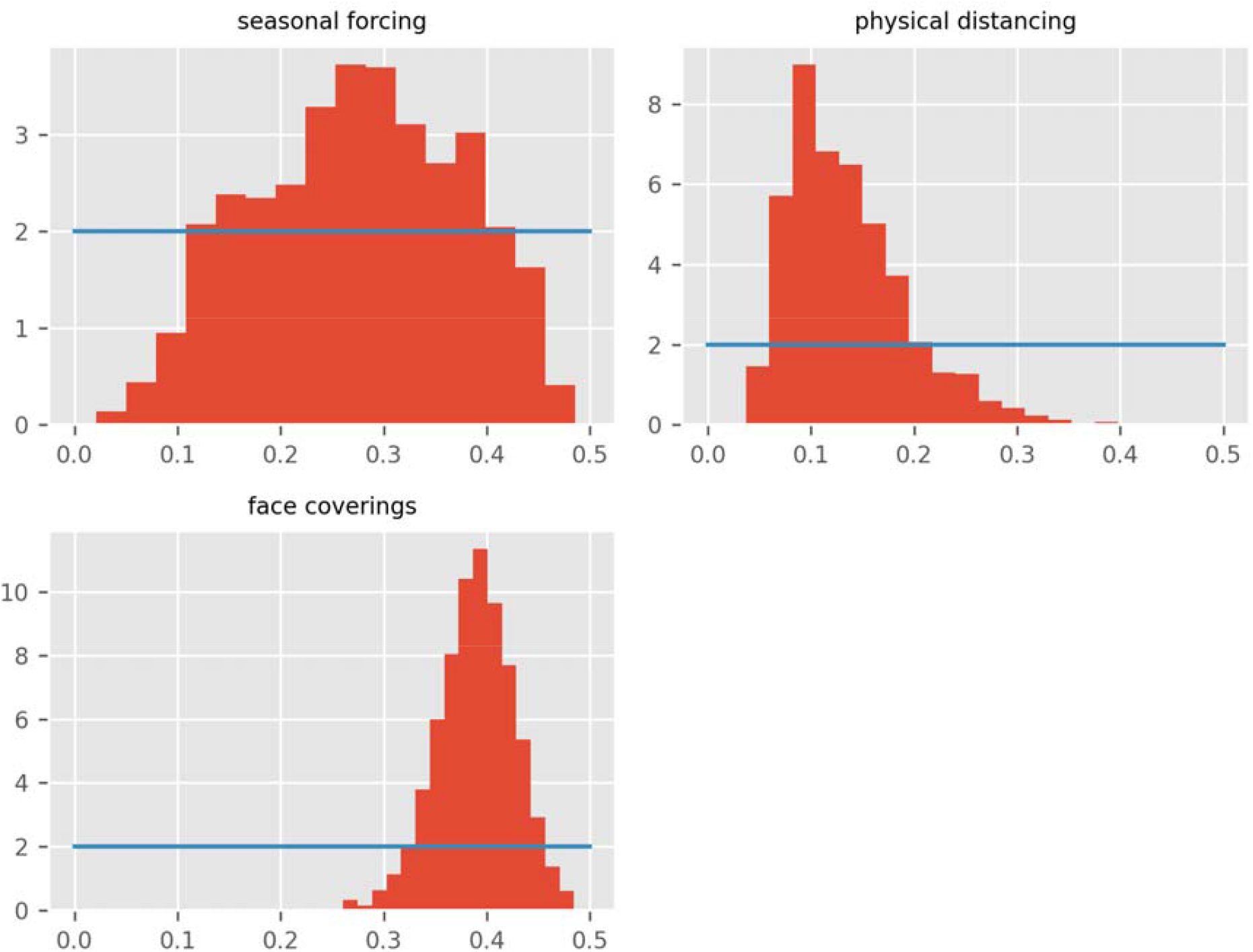
Posterior density histograms for key state-wide epidemiological parameters from accepted model runs. Red histograms, model posterior estimates; blue lines, prior distributions for same parameters (all are uniform or truncated normal distributions).

**Figure 6.**
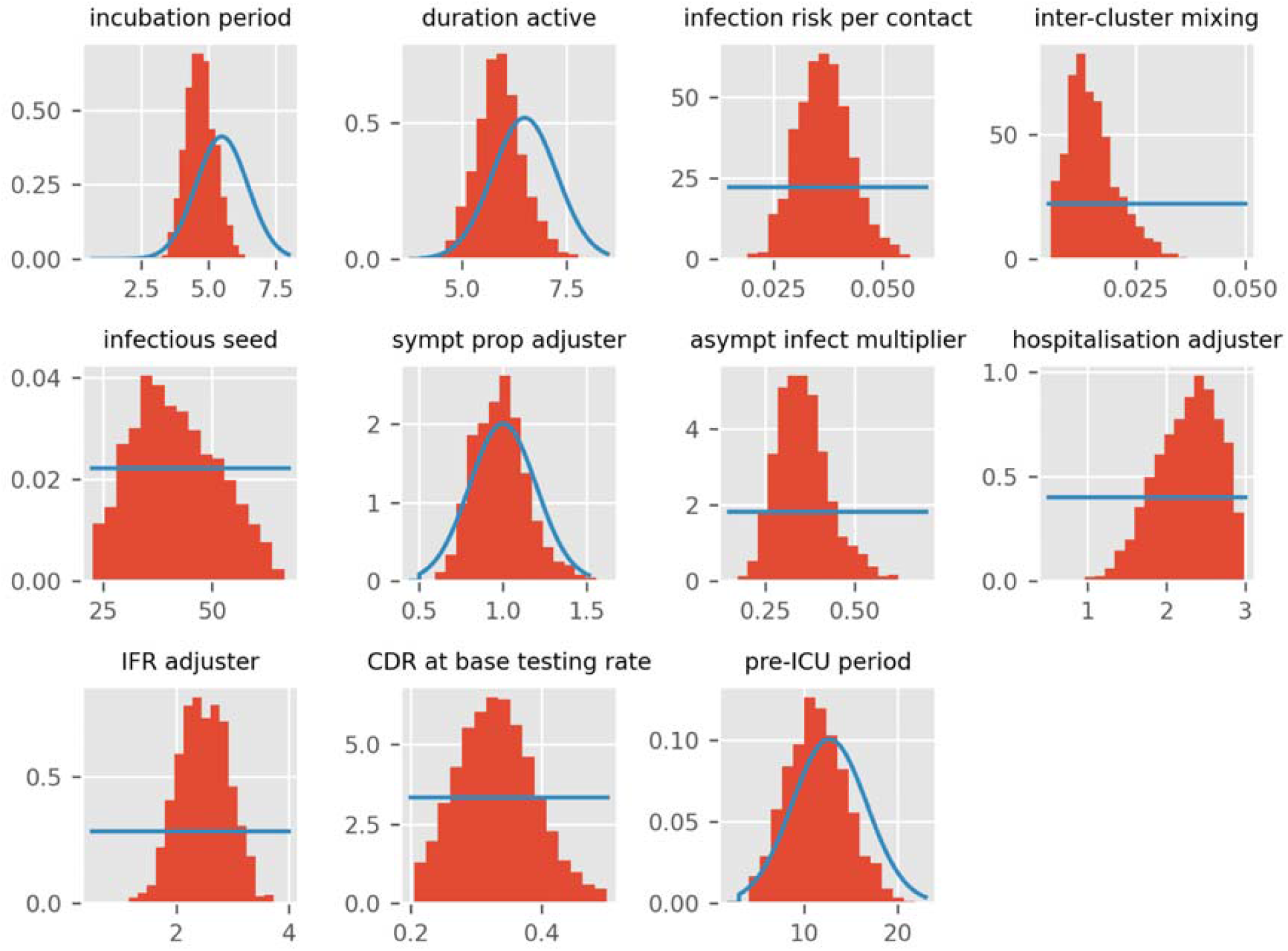
Posterior histograms for other state-wide epidemiological parameters. Red histograms, model posterior estimates; blue lines, prior distributions for same parameters (all are uniform or truncated normal distributions).

**Figure 7.**
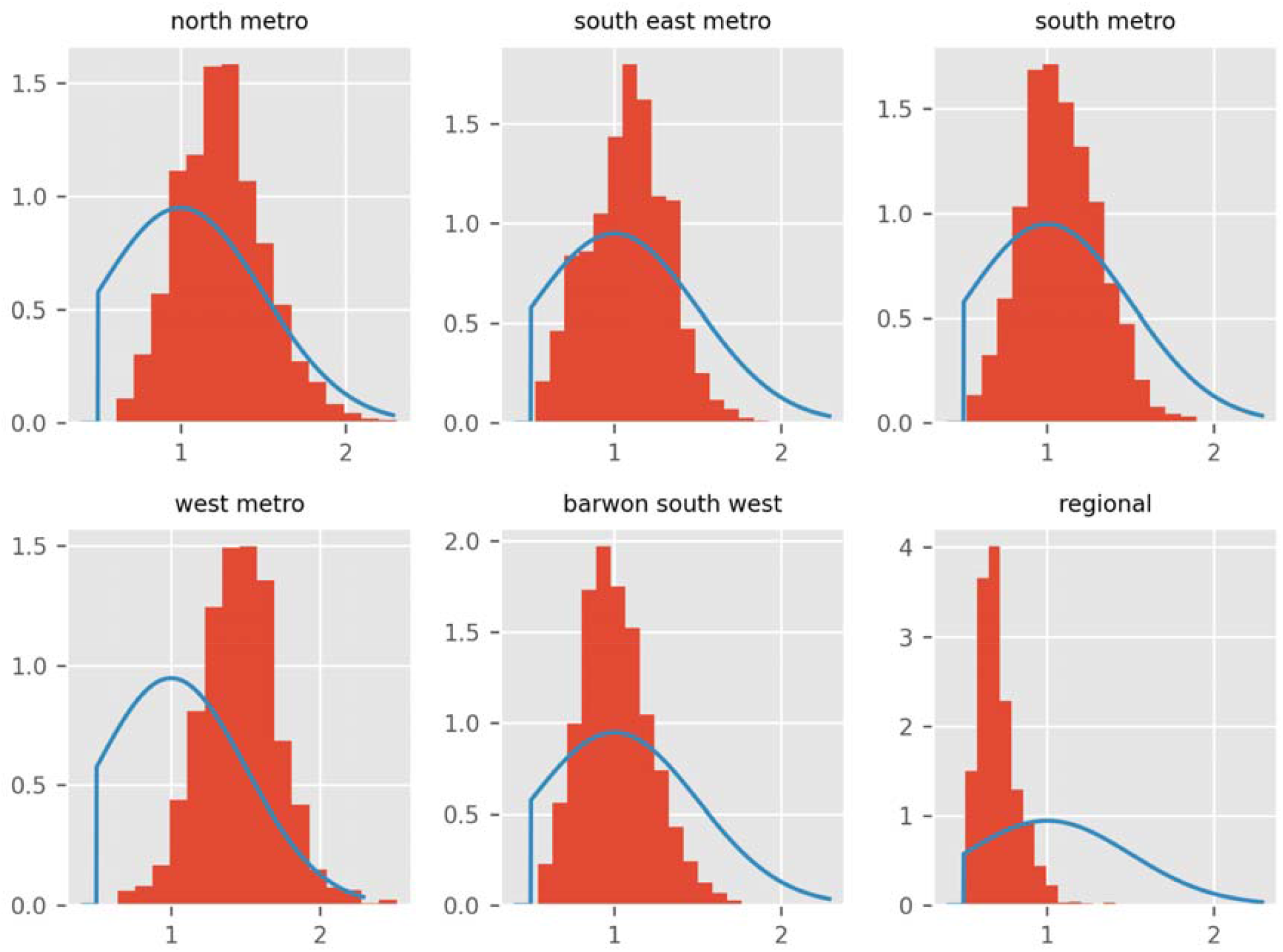
Posterior histograms for cluster-specific contact rate modifier parameters. Red histograms, model posterior estimates; blue lines, prior distributions for same parameters (all are uniform or truncated normal distributions).

**Figure 8.**
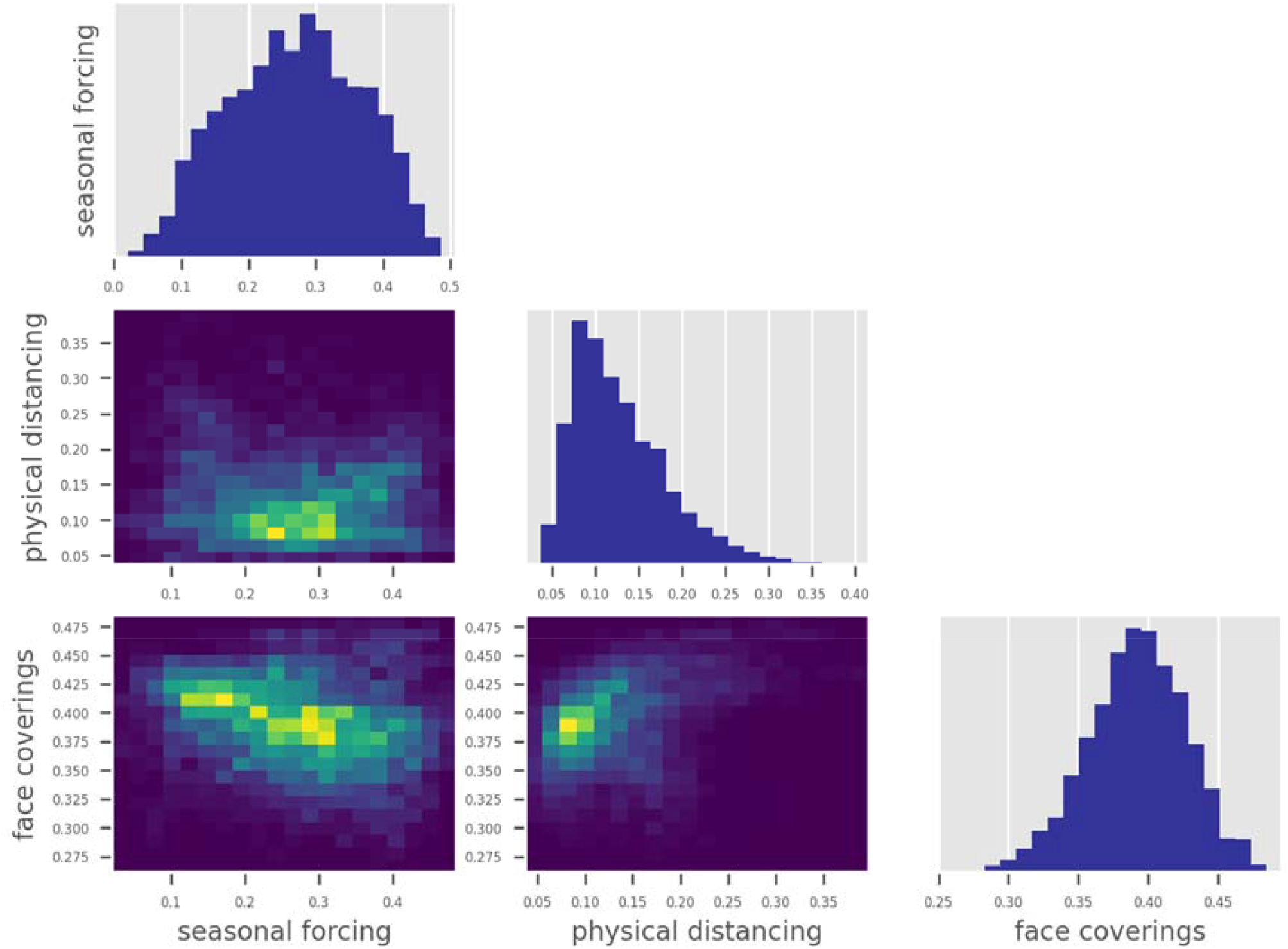
Correlation matrix for key state-wide epidemiological parameters.

**Figure 9.**
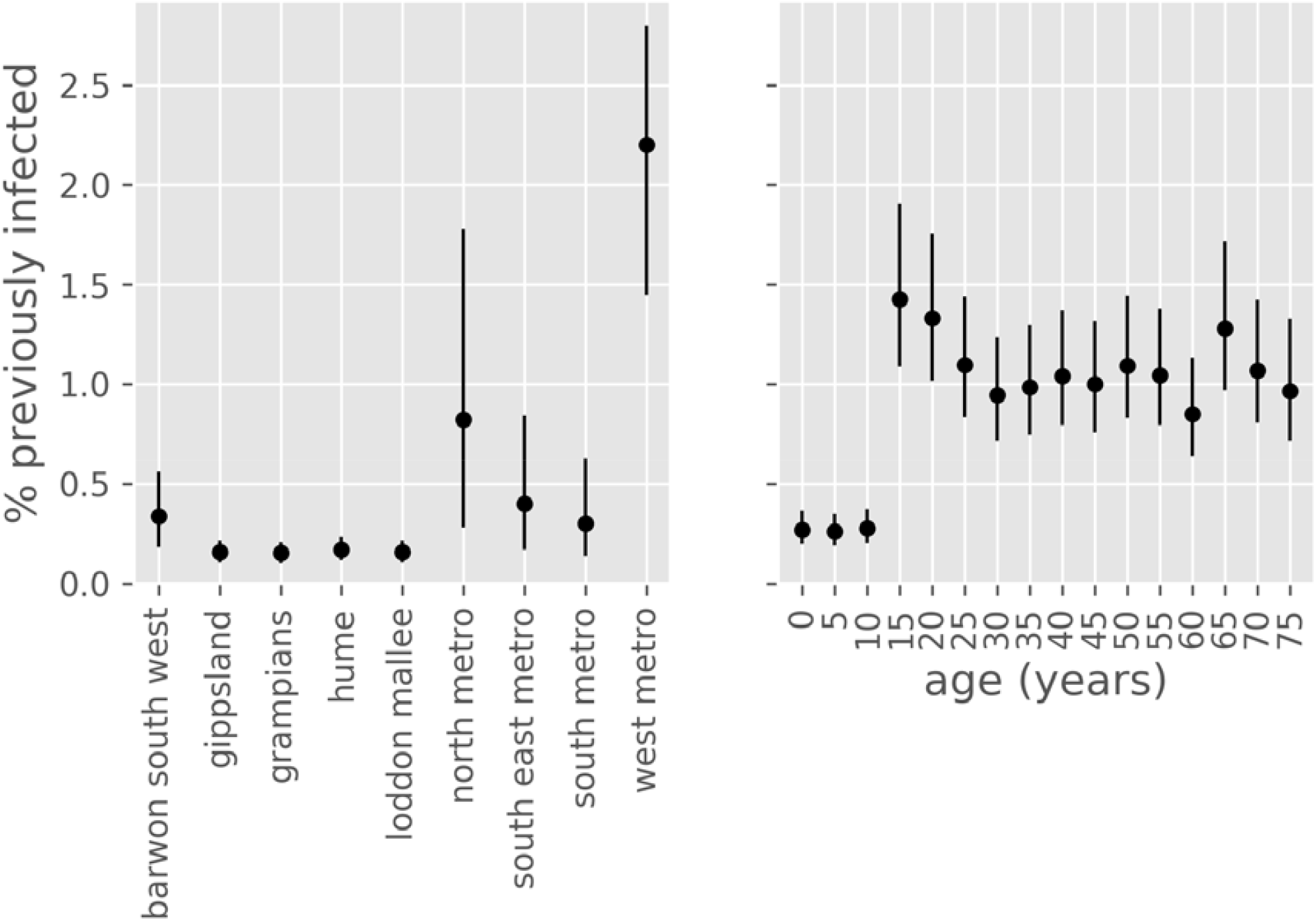
Estimated proportion of population recovered from COVID-19 at 1st October 2020, by age group and health service cluster. Point estimates with associated 50% credible intervals. Values are negligibly different from attack rates, except that deaths are excluded from the denominator.

### Parameter estimation

The posterior estimates of model calibration parameters are presented in Table 1. Several epidemiological parameters with good evidence from international studies showed posteriors that were consistent with prior beliefs. This prevented overfitting, reduced the degree of freedom and provided better estimates of key free parameters including the effect of time-varying processes, allowing insights into the dynamics of the epidemic. The unadjusted risk of transmission per contact (specifically the risk of transmission per contact between a susceptible person aged 15-64 years and a symptomatic infectious person not in isolation) was estimated at 2-5%. This needed to be adjusted for each cluster modelled, with the modifiers applied to the metropolitan clusters reaching values up to double that for the regional clusters (other than Barwon South West). The extent of mixing between geographical patches was low, with around 1-2% of the total force of infection contributed by regions other than the index patch.

Estimates of the incubation period, the infectious period, the period prior to ICU admission and the duration in ICU were similar to our prior estimates derived from the literature. Likewise, the estimated proportion of incident cases resulting in symptomatic disease was similar to our prior estimate. However, the risk of hospitalisation (and hence ICU admission) and of death given infection were considerably greater than our age-specific prior estimates obtained from the literature. This likely reflects higher rates of exposure and infection in population groups at particularly high risk of adverse outcomes, including residents of aged care facilities.

The case detection rate associated with a testing rate of one test per 1,000 population per day was estimated at 33.0% (95%CI, 22.4-45.3), such that peak rates of detection of symptomatic infections were estimated at greater than 60%.

To understand the reasons behind the epidemic curve plateauing at the start of August and beginning to decline thereafter, we were particularly interested in parameters governing the effect of time-varying processes. We estimated that physical distancing behaviours and face coverings were both important in achieving control of Victoria’s second wave, with face coverings estimated to have reduced transmission and infection risk by around 31 to 46%, while fitting to data provided little information on the effect of seasonal forcing (around 9 to 45%). Physical distancing behaviour was estimated to have reduced risk of transmission/infection by around 6 to 28%, although the smaller changes in reported adherence to this intervention (Supplemental Figure 5) meant that this had a lesser impact on the epidemic profile. For the behavioural changes in particular, the posterior probability density was substantially more informative than the prior and had negligible density around the value of zero, consistent with an effect of each of these interventions in reversing the epidemic trajectory. Additionally, the posterior probabilities of the parameters were only moderately collinear (Figure 8), supporting independent effects for each process.

### Counterfactual scenarios

Figure 10 presents four counterfactual scenarios in addition to the baseline scenario. The effect of re-opening schools from 9th July (the date that stage 3 restrictions were imposed) was projected to be modest, with daily case rates peaking around 200 higher than under baseline conditions, but with the epidemic profile otherwise broadly similar. The effect of not mandating face coverings was projected to be dramatic, with case numbers in the thousands for several months under the counterfactual of face coverings usage remaining at the baseline level of 13.0%. Returning to full mobility from 9th July resulted in a similarly poorly controlled epidemic, under the assumption that face coverings usage could not then have reached the baseline estimate of >90% compliance in all workplaces and other locations if industries such as hospitality were fully re-opened. An epidemic unmitigated by any movement and behavioural restrictions was projected to substantially overwhelm expanded ICU capacity.

**Figure 10.**
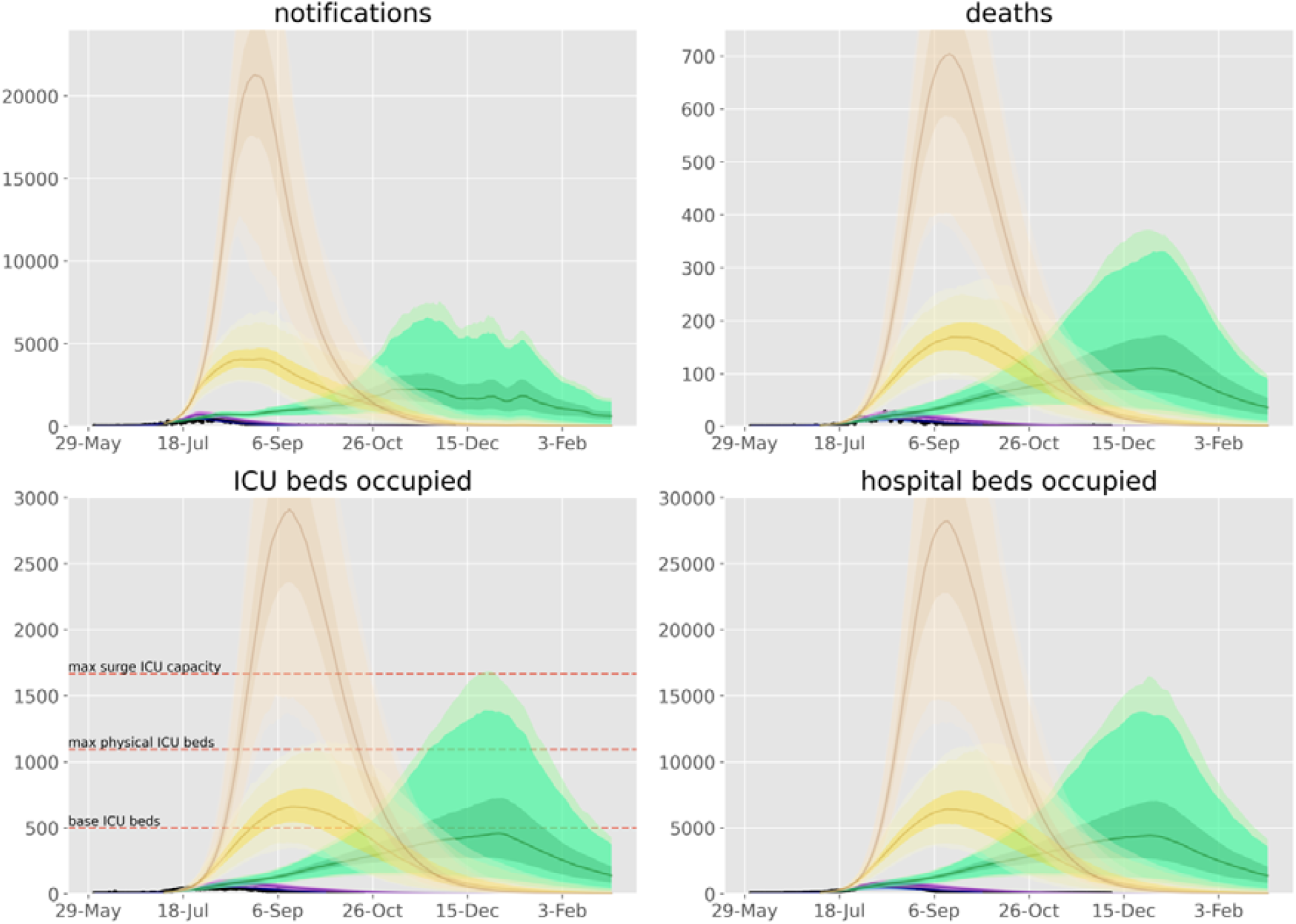
Counterfactual scenarios compared against baseline calibration and data. Scenarios are: purple, schools re-opened from 7th July; green, face coverings not mandated (on 23rd July); yellow, work, schools and other locations mobility return to baseline levels from 7th July with 60% face coverings compliance; brown, return to normal mobility with baseline use of face coverings. Data (black dots), median modelled estimates (lines), shaded areas 25th to 75th centile (darkest shading), 2.5th to 97.5th centile (intermediate shading depth) and 1st to 99th centile (faintest shading) of each indicator for each scenario. 7th July chosen as the date that stage 3 restrictions were imposed. We considered that full compliance with mandatory face coverings would be impractical if workplaces and other locations returned to full capacity (for example, if hospitality was fully re-opened, patrons would not wear masks in all other locations). Base and surge ICU capacity for Victoria presented on lower left panel.^30^

## Discussion

We found that the improvement in Victoria’s second wave of COVID-19 cases could be well captured in our transmission model through a combination of time-variant processes that included: testing rates, population mobility, use of face coverings, physical distancing and seasonal forcing. The lower rates of COVID-19 observed in regional clusters were captured with the introduction of the infectious seed through the metropolitan clusters only, without needing to unrealistically manipulate the risk of transmission by cluster. Risk of infection in metropolitan areas was estimated to be up to double that of regional areas, consistent with international findings of a moderate correlation between population density and epidemic severity.^5^ Although Barwon South West showed transmission rates that were more comparable to metropolitan Melbourne, this region includes Victoria’s second largest city of Geelong. Interaction between populations of different clusters was low in the context of significant restrictions on movement between regions. Each of the time-varying processes modelled appeared to be important to the observed dynamics, with both face coverings and behavioural changes associated with a significant reduction in transmission risk per contact. However, face coverings had a considerably greater effect on reversing the epidemic, which was observable due to the sharp transition in the extent of their use when they were mandated.

Victoria’s second wave of cases was dramatically different from its first autumn wave, which was driven by importations and during which time the effective reproduction number was consistently estimated to be below one.^14,15^ Victoria’s second wave was initiated by quarantine escape, from which widespread community transmission soon followed. Progressively more extensive lockdown measures were then implemented, with local targeting of specific residential blocks and then postcodes, which were insufficient to reverse the epidemic trajectory.

As noted previously, stage 3 restrictions were associated with a reduction in the effective reproduction number,^16^ although significant case rates persisted throughout July, and further reductions in mobility were observed with stage 4. An agent-based model with detailed social networks consideration of multiple intervention types and without geographical structure was calibrated to the Victorian epidemic.^17^ This model emphasised the importance of associations between individuals who would not otherwise be in regular contact to the epidemic. By contrast to previous work, our model captures the temporal and spatial implementation of the policy changes in Victoria to allow inference of the effect of each intervention. As concern increased that epidemic control had not been achieved over the course of July, policy changed rapidly in an attempt to bring the epidemic under control. Testing numbers increased following a nadir in early June and lockdown measures were implemented differently in twelve Melbourne postcodes, the remaining postcodes of Greater Melbourne, Mitchell Shire (immediately north of Greater Melbourne) and the remainder of regional Victoria. We captured these complicated geographical patterns of restriction by scaling our mixing matrices using Google mobility data, which are available at the LGA level for Victoria. School closure and face covering policy changes were captured according to the dates of policy changes.

Most of the inferred parameters were consistent with previous evidence, including a potentially important effect of seasonal forcing in terms of the absolute reduction in virus transmissibility.^18^ The minimal information provided on seasonal forcing is likely attributable to our simulation period spanning less than four months and so covering a small proportion of the cycling period, such that the effect could represent other secular changes during the period modelled. Sensitivity analysis (not presented) with seasonal forcing set to zero throughout the simulations made a negligible difference to the estimates for the other parameters of interest. The effect of face coverings was greater than is typically estimated at the individual level,^19^ but is consistent with the dominant importance of the respiratory route to transmission.^20^ The finding was also not unexpected given the marked shift in population use of face coverings at this time and the timing of the policy change in late July relative to the dramatic reversal in case numbers occurring around one week later. The significant estimated effect of behavioural changes suggests that reductions in interpersonal associations (macro-distancing) alone were insufficient to achieve the dramatic reversal in the epidemic trajectory observed. However, the Google mobility functions used to capture macro-distancing simulated falls in attendance at workplaces and other non-household locations to considerably below baseline values in several clusters (Figure 1), emphasising their importance. The dramatic effect of each of these interventions on the epidemic trajectory is partly attributable to our implementation of these processes as applying to both the infectious cases and the exposed individual. This approach is analogous to simulating the use of bed-nets for malaria control, where the overall effect of the intervention is quadratic, as it affects both the disease vector and the infection reservoir.^21^

Despite the complexity of our model, it is inevitably a simplification of reality. Although we assumed that asymptomatic cases were undetectable, this was addressed by varying two epidemiological parameters pertaining to these patients, and the posterior estimate of the infectiousness of asymptomatic cases suggested around threefold lower infectiousness per unit time. Our findings relating to the impact of time-varying interventions could be proxies for other effects. For example, although we considered that the effectiveness of case detection scaled with the number of tests performed, changes in the effectiveness of tracing, testing and isolation activities during the course of the epidemic wave may also have been important.

Victoria’s second wave is known to have had particularly dramatic effects on residents of aged care facilities and health care workers, which we did not explicitly capture except by varying parameters relating to disease severity. Because of this, it was necessary to scale international estimates of the age-specific infection fatality rate around two- to three-fold. Although this factor seems extreme, age-specific infection fatality rate estimates increase dramatically with age,^7^ and it should be noted that the age-specific infection fatality rate parameters we used increase up to three-fold with each successive decade of age.^6^ Therefore, an age distribution of the infected population that is one decade higher than that simulated would be expected to have a comparable effect. Our age-specific estimates of the risk of hospitalisation given symptomatic COVID-19 do not fully capture the consideration that hospital admission is driven by factors other than disease severity, including infection control and workforce capacity and staff isolation requirements in residential aged care facilities, which were particularly important to this epidemic wave.

With the state’s explicit objective of achieving no community transmission in Victoria (and therefore across Australia) within a few months,^22,23^ our findings emphasise that multiple interacting components of the public health interventions were required to achieve this within the modelled period.^24,25^ Consistent with findings from elsewhere,^26–28^ without reductions in contacts outside the home and mandating the use of masks, there would have been no reasonable prospect of driving transmission to zero within a time period tolerable to the community, given the starting point of the epidemiological situation in late July. The small effect of school closures was also consistent with findings from overseas,^25,29^ although if schools had remained open throughout the epidemic wave, some additional weeks would likely have been required for transmission to decline to the point that elimination was an immediate prospect. Nonetheless, it is encouraging that in a low transmission scenario, school closures are likely not necessary to gain control in the presence of other effective population-level restrictions, including masks.

In conclusion, we found that Victoria’s major second wave of COVID-19 was brought under control through a combination of policy interventions that were synergistic and together contributed substantially to the dramatic reversal in the observed epidemic trajectory. In particular the considerable individual-level effect of face coverings was critical to achieving epidemic control, and so should be a cornerstone of any public health response given the much lesser inconvenience associated with their use compared to restrictions on mobility. Rates of hospitalisation and death were higher than anticipated given international estimates of parameters pertaining to these quantities, likely reflecting the concentration of the epidemic in high-risk groups, particularly residents of aged care facilities. As vaccination is rolled out as a more targeted intervention, protection of high-risk settings, including aged care will be critical, particularly in regions with low or negligible population immunity that remain at risk of explosive epidemics.

## Data Availability

All code used in the simulations presented is available at https://github.com/monash-emu/AuTuMN. The calibration data were provided in confidence by the Victorian Department of Health and Human Services, but are numerically very similar to those already in the public domain.

https://github.com/monash-emu/summer

## Author declarations

BS and ACC wish to emphasise their important statutory roles during Victoria’s pandemic response in 2020, as Chief Health Office and acting Chief Health Officer respectively. MJL and GWD were also employed by DHHS during 2020. JMT provided regular advice to DHHS during this time as an independent advisor.

## Acknowledgements

We gratefully acknowledge the support and advice of staff of the Victorian Department of Health and Human Services (now the Victorian Department of Health) for provision of data and assistance with its interpretation. We thank A/Prof Nicholas Golding for providing the micro-distancing compliance factors.

## Contributor statement

JMT and RR constructed the model. GD, MJL and JN assisted with provision of data. JMT wrote the first draft of the manuscript, which was then revised with input from all authors.

