## Supplementary Appendix for "Understanding how Victoria, Australia gained control of its second COVID-19 wave"

Epidemiological Modelling Unit,

School of Public Health and Preventive Medicine

Australian Tuberculosis Modelling Network

**5 Between cluster mixing** **17**

**6 Model initialisation** **20**

**7 Parameters** **20**

**10 Likelihood function** **29**

**11 Ordinary differential equations** **29**

**12 Supplemental Output Figures** **34**

### 1 Base model construction

#### 1.1 Platform for infectious disease dynamics simulation

We developed a deterministic compartmental model of COVID-19 transmission using the AuTuMN platform, publicly available at https://github.com/monash-emu/AuTuMN/. Our repository allows for the rapid and robust creation and stratification of models of infectious disease epidemiology and includes pluggable modules to simulate heterogeneous population mixing, demographic processes, multiple circulating pathogen strains, repeated stratification and other dynamics relevant to infectious disease transmission. The platform was created to simulate TB dynamics, being an infectious disease whose epidemiology differs markedly by setting, such that considerable flexibility is desirable [1]. We have progressively developed the structures of our platform over recent years, and further adapted it to be sufficiently flexible to permit simulation of other infectious diseases for the purpose of this project.

#### 1.2 Base COVID-19 model

Using the base framework of an SEIR model (susceptible, exposed, infectious, removed), we split the exposed and infectious compartments into two sequential compartments each (SEEIIR). The two sequential exposed compartments represent the non-infectious and infectious phases of the incubation period, with the latter representing the "presymptomatic" phase such that infectiousness occurs during three of the six sequential phases. For this reason, "active" is a more accurate term for the two sequential "I" compartments and is preferred henceforward. The two infectious compartments represent early and late phases of active disease, during which symptoms occur if the disease episode is symptomatic, and allow explicit representation of notification, case isolation, hospitalisation and admission to ICU. The "active" compartment also includes some persons who remain asymptomatic throughout their disease episode, such that these compartments do not map directly to either persons who are infectious or those who are symptomatic (Figure 1).

The latently infected and infectious presymptomatic periods together comprise the incubation period, with the incubation period and the proportion of this period for which patients are infectious defined by input parameters described below. In general, two sequential compartments can be used to form a gamma-distributed profile of transition to infectiousness following exposure if the progression rates for these two compartments are equal, although in implementing this model the relative sojourn times in the two sequential compartments usually differed. Nevertheless, the profiles implemented are broadly consistent with the empirically observed log-normal distribution of individual incubation periods [2].

The transition from early active to late active represents the point at which patients are detected (for those persons for whom detection does eventually occur) and isolation then occurs from this point forward (i.e. applies during the late disease phase only, see Section 2). This transition point is also intended to represent the point of admission to hospital or transition from hospital ward to intensive care for patients for whom this occurs (see Section 1.4).

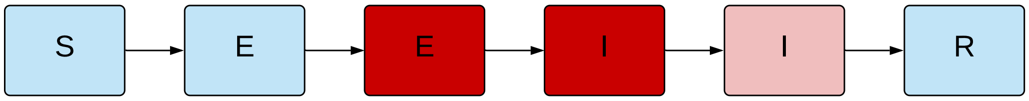

**Figure 1 – Unstratified compartmental model structure.** S = susceptible, E = exposed, I = active, R = recovered/removed. Depth of pink/red shading indicates the infectiousness of the compartment.

##### *1.3 Age stratification*

All compartments of this base compartmental structure were stratified by age into five-year bands from 0-4 years of age through to 70-74 years of age, with the final age group being those aged 75 years and older. Heterogeneous baseline contact patterns by age were incorporated using age-specific contact rates estimated by Prem et al. 2017 [3], who combined survey response data with information on national demographic characteristics to produce age-structured mixing matrices with these age groupings. These are then modified by non-pharmaceutical interventions as described in Section 3. Our modelled age groups were chosen to match these mixing matrices. The automatic demographic features of AuTuMN that can be used to simulate births, ageing and deaths were not implemented, because the issues considered pertain to the short- to medium-term and the immediate implementation of control strategies, for which population demographics are less relevant.

##### 1.4 Clinical stratification

The age-stratified late exposed/incubation and both the early and late active disease compartments were further stratified into five "clinical" categories: *1)* asymptomatic, *2)* symptomatic ambulatory, never detected, *3)* symptomatic ambulatory, ever detected, *4)* ever hospitalised, never critical and *5)* ever critically unwell (Figure 2). The proportion of new infectious persons entering stratum 1 (asymptomatic) is age-dependent (as described in Table 5). The proportion of symptomatic patients (strata 2 to 5) ever detected (strata 3 to 5) is set through a parameter that represents the time-varying proportion of all symptomatic patients who are ever detected (the case detection rate, see Section 2). Of those ever symptomatic (strata 2 to 5), a time-constant but age-specific proportion is considered to be hospitalised (entering strata 4 or 5). Of those hospitalised (entering strata 4 or 5), a fixed proportion was considered to be critically unwell (entering stratum 5, Figure 3).

##### 1.5 Hospitalisation

For COVID-19 patients who are admitted to hospital, the sojourn time in the early and late active compartments is modified, superseding the default values of the sojourn times for these compartments, as indicated in Table 4. The point of admission to hospital is considered to be the transition from early to late active disease, such that the sojourn time in the late disease represents the period of time admitted to hospital. For patients admitted to ICU, admission to ICU occurs at this same transition point. For this group, the period of time hospitalised prior to ICU admission is estimated as a proportion of the early active period, such that the early active period represents both the period ambulatory in the community and the period in hospital prior to ICU admission.

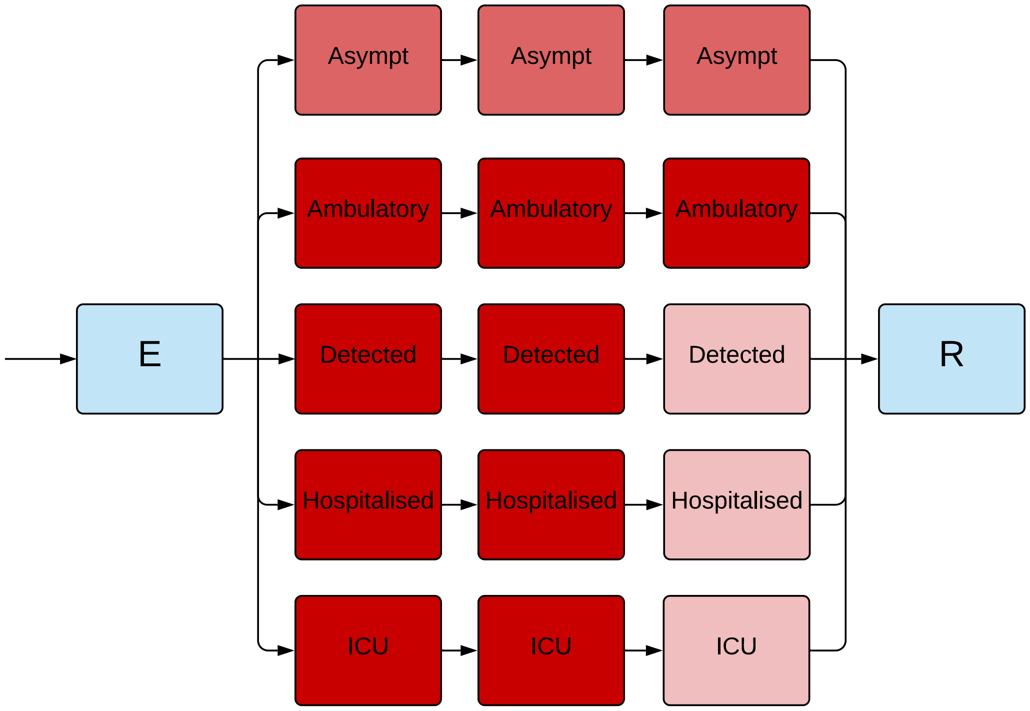

**Figure 2 – Illustration of the implementation of the clinical stratification.** Depth of pink/red shading indicates the infectiousness of the compartment. Typical parameter values presented, although the infectiousness of asymptomatic persons is varied in calibration.

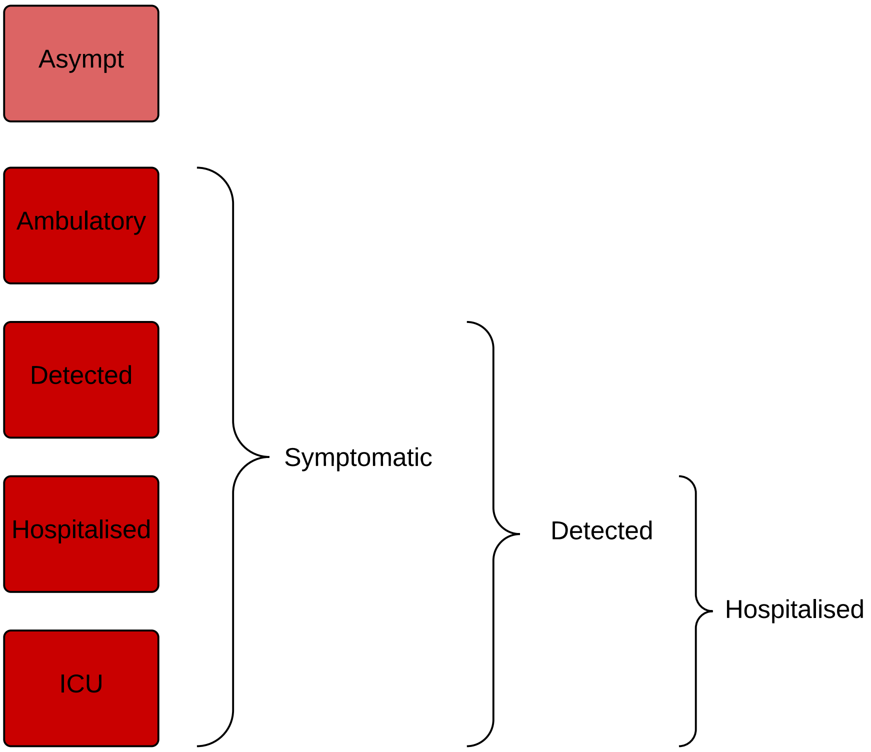

**Figure 3 – Illustration of the rationale for the clinical stratification.**

##### 1.6 Infectiousness

Asymptomatic persons are assumed to be less infectious per unit time active than symptomatic persons not undergoing case isolation (typically by around 50%, although this is varied in calibration/uncertainty analysis). Infectiousness is also decreased for persons who have been detected to reflect case isolation, and for those admitted to hospital or ICU to reflect infection control procedures (by 80% for both groups). Presymptomatic individuals are presumed to have equivalent infectiousness to those with early active COVID-19.

##### 1.7 Application of COVID-19-related death

Age-specific infection fatality rates (IFRs) were applied and distributed across strata 4 and 5, with no deaths typically applied to the first three strata. A ceiling of 50% is set on the proportion of those admitted to ICU (entering stratum 5) who die. If the infection fatality rate is greater than this ceiling, the proportion of critically unwell persons dying was set to 50%, with the remainder of the infection fatality rate then applied to the hospitalised proportion. Otherwise, if the infection fatality rate is less than half of the absolute proportion of persons critically unwell, the infection fatality rate is applied entirely through stratum 5 (such that the proportion of critically unwell persons dying in that age group becomes $<$50% and the proportion of stratum 4 dying is set to zero). In the event that the infection fatality rate for an age group is greater than the total proportion hospitalised (which is unusual, but could occur for the oldest age group under certain parameter configurations), the remaining deaths are assigned to the asymptomatic stratum. This approach was adopted for computational ease and is valid because the duration active for persons entering this stratum is the same as for the other non-hospitalised strata, such that the dynamics are identical to assigning the deaths to any of the first three strata. We used the age-specific IFRs previously estimated from age-specific death data from 45 countries and results from national-level seroprevalence surveys [4] as indicated in Table 5. We allowed IFRs to vary around the previously published point estimates in order to incorporate uncertainty and to allow the IFRs to differ from the settings in which they were estimated (see Calibration section).

| Clinical stratum | Stratum name | Pre-symptomatic | Early | Late |
|---|---|---|---|---|
| 1 | Asymptomatic | 0.5 | 0.5 | 0.5 |
| 2 | Symptomatic ambulatory never detected | 1 | 1 | 1 |
| 3 | Symptomatic ambulatory ever detected | 1 | 1 | 0.2 |
| 4 | Hospitalised never critical | 1 | 1 | 0.2 |
| 5 | Ever critically unwell | 1 | 1 | 0.2 |

**Table 1 – Illustration of the relative infectiousness of disease compartments by clinical stratification and stage of infection.** Typical parameter values displayed.

##### 1.8 Seasonal forcing

Seasonal forcing is implemented through a simple sinusoidal function that is multiplied by the contact probability of the form:

$$\begin{aligned} contact\ probability(time) = cos((time - peak\,time) \times 2 \times \pi \div 365) \\ \times forcing \div 2 \\ + average\,contact\,rate, \end{aligned} \quad (1)$$

such that *time* is the time in days from the 31st December 2019 and *peak time* is the date of the winter solstice (=173). *forcing* is the relative magnitude of peaks compared to troughs in the probability of transmission per contact induced by seasonal forcing. This notation is consistent with that of others [5], except that previous similar formulas present the minimum (summer) contact rate added to the seasonal variation, whereas we consider the average (or equinox) contact rate to be a more intuitive parameter (i.e. $average\ contact\ rate = minimum\ contact\ rate + forcing\ \div\ 2$ [5]).

#### 2 Case detection

##### 2.1 General approach

We calculate a time-varying case detection rate, being the proportion of all symptomatic cases (clinical strata 2 to 5) that are detected (clinical strata 3 to 5). This proportion is informed by the number of tests performed using the following formula:

$$CDR(time) = 1 - e^{-shape \times tests(time)}$$

*time* is the time in days from the 31st December 2019 and *tests(time)* is the number of tests per capita done on that date. To determine the value of the shape parameter, we solve this equation based on the assumption that a certain daily testing rate *tests(t)* is associated with a certain *CDR(t)*. Solving for *shape* yields:

$$shape = \frac{-log(1 - CDR(t))}{tests(t)}$$

That is, if it is assumed that a certain daily per capita testing rate is associated with a certain proportion of symptomatic cases detected, we can determine *shape*. As this relationship is not well understood and unlikely to be consistent across all settings, we vary the *CDR* that is associated with a certain per capita testing rate during uncertainty/calibration. Given that the *CDR* value can be varied widely, the purpose of this is to incorporate changes in the case detection rate that reflect the empirical historical profile of changes in testing capacity over time.

##### 2.2 Testing data

Statewide daily testing data by date of test were provided by DHHS and applied to all health system clusters to provide a broad profile of the variation in testing capacity over time, including the lower testing numbers in early June compared to at the peak of the epidemic (Figure 4). Data sparseness precluded us from implementing separate functions for each individual health service cluster. For this application to Victoria, the case detection proportion corresponding to a per capita rate of testing of one test per thousand population per day was varied as a calibration parameter in creating the time-varying case detection proportion function. Note that testing rates were typically considerably higher than one per thousand per day during the period modelled, such that the actual modelled case detection proportion is considerably higher than the case detection calibration parameter for most of the simulation period.

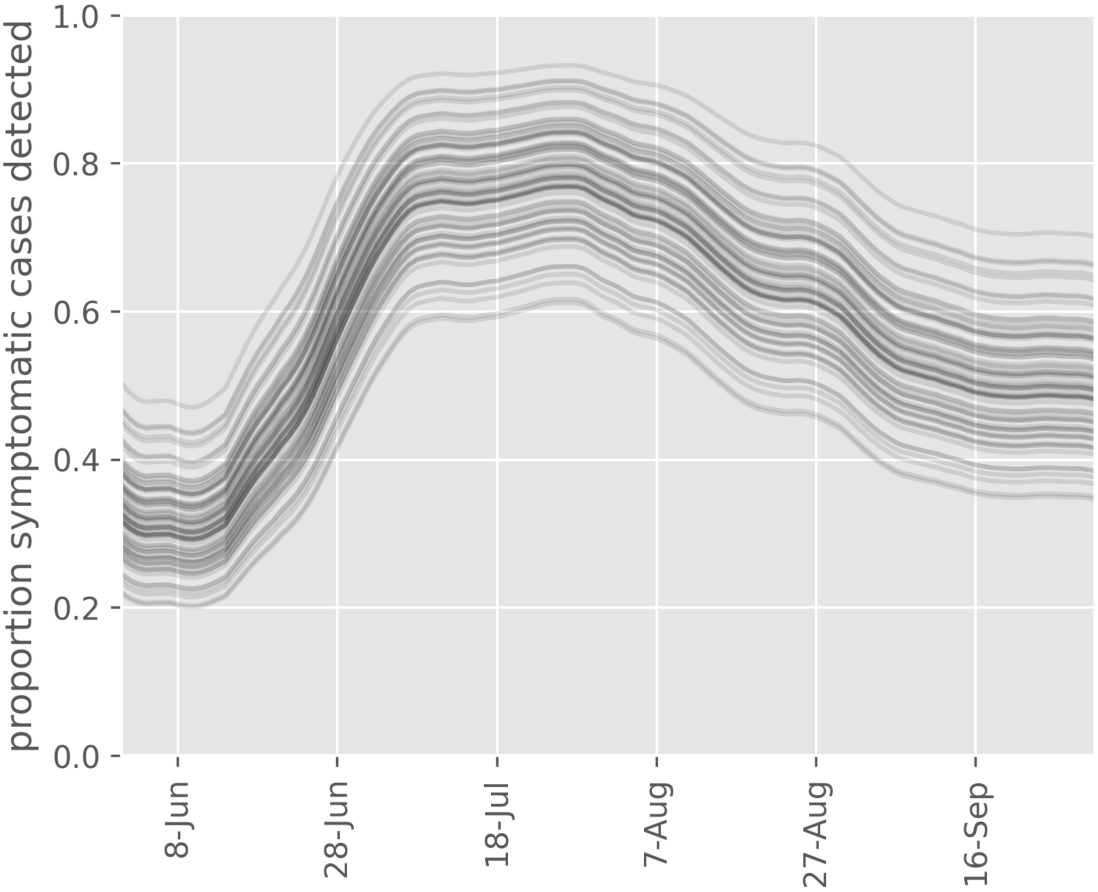

**Figure 4 – Example case detection rate curves.** 70 parameters for the case detection rate at a given daily per capita testing rate were sampled at random from accepted values to produce this graph.

### 3 Implementation of non-pharmaceutical interventions

A major part of the rationale for the development of this model was to capture the past impact of non-pharmaceutical interventions (NPIs) and produce future scenarios projections with the implementation or release of such interventions.

#### 3.1 Isolation and quarantine

For persons who are identified with symptomatic disease and enter clinical stratum 3, self-isolation is assumed to occur and their infectiousness is modified as described above. The proportion of ambulatory symptomatic persons effectively identified through the public health response by any means is determined by the case detection rate as described above.

#### 3.2 Community quarantine or "lockdown" measures

For all NPIs relating to reduction of human mobility or "lockdown" (i.e. all NPIs other than isolation and quarantine), these interventions are implemented through dynamic adjustments to the age-assortative mixing matrix. The baseline mixing matrices of Prem et al. [3] are synthetic and do not represent direct observations or reports from surveys (in the case of the 144 countries to which they were extrapolated from observations in the eight "POLYMOD" countries of Western Europe). Although synthetic, the matrices are contextualised to national demographic information, including country-specific data that include household size, workforce participation and school enrolment. Further, the matrices presented are easily machine-readable and appear to be plausible representations of contact structures within these countries.

The matrices also have the major advantage of allowing for disaggregation of total contact rates by location, i.e. home, work, school and other locations. This disaggregation allows for the simulation of various NPIs in the local context by dynamically varying the contribution of each location to reflect the historical implementation of the interventions.

The corresponding mixing matrix (denoted $C_0$) is presented using the standard convention that a row represents the average number of age-specific contacts per day for a contact recipient of a given age-group. In other words, the element $C_{0i,j}$ is the average number of contacts per day that an individual of age-group

$_j$ makes with individuals of age-group $_i$.

This matrix results from the summation of the four location-specific contact matrices provided by Prem et al.: $C_0 = C_H + C_S + C_W + C_L$, where $C_H$, $C_S$, $C_W$ and $C_L$ are the age-specific contact matrices associated with households, schools, workplaces and other locations, respectively.

In our model, the contributions of the matrices $C_S$, $C_W$ and $C_L$ vary with time such that the input contact matrix can be written:

$$C(t) = C_H + s(t)^2 C_S + w(t)^2 C_W + l(t)^2 C_L$$

The modifying functions are each squared to capture the effect of the mobility changes on both the infector and the infectee in any given interaction that could potentially result in transmission. The modifying functions incorporate both macro-distancing and microdistancing effects, depending on the location.

##### 3.3 School closures/re-openings

Reduced attendance at schools is represented through the function $s(t)$, which represents the proportion of all school students currently attending on-site teaching. If schools are fully closed, $s(t) = 0$ and $C_S$ does not contribute to the overall mixing matrix $C(t)$. $s(t)$ is calculated through a series of estimates of the proportion of students attending schools, to which a smoothed step function is fitted. Note that the dramatic changes in this contribution to the mixing matrix with school closures/re-openings is a more marked change than is seen with the simulation of policy changes in workplaces and other locations (which are determined by empiric data and so do not vary so abruptly and do not fall to zero).

##### 3.4 Workplace closures

Workplace closures are represented by quadratically reducing the contribution of workplace contacts to the total mixing matrix over time. This is achieved through the scaling term $w(t)^2$ which modifies the contribution of $C_W$ to the overall mixing matrix $C(t)$. The profile of the function $w(t)$ is set by fitting a polynomial spline function to Google mobility data for workplace attendance (Table 2).

##### *3.5 Community-wide movement restriction*

Community-wide movement restriction (or "lockdown") measures are represented by proportionally reducing the contribution of the other locations contacts to the total mixing matrix over time. This is achieved through the scaling term $l(t)^2$ which modifies the contribution of $C_L$ to the overall mixing matrix $C(t)$. The profile of the function $l(t)$ is set by fitting a polynomial spline function to an average of Google mobility data for various locations, as indicated in Table 2.

##### *3.6 Household contacts*

The contribution of household contacts to the overall mixing matrix $C(t)$ is fixed over time. Although Google provides mobility estimates for residential contacts, the nature of these data are different from those for each of the other Google mobility types in that they represent the time spent in that location rather than the duration. The daily frequency with which people attend their residence is likely to be close to one and we considered that household members likely have a daily opportunity for infection with each other household member. Therefore, we did not implement a function to scale the contribution of household contacts to the mixing matrix with time.

| **Prem "location"** | **Approach** | **Google mobility types** |
|---|---|---|
| School | Policy response | Not applicable |
| Household | Constant | Not applicable |
| Workplace | Google mobility | Workplace |
| Other locations | Google mobility | Unweighted average of:<br>• Retail and recreation<br>• Grocery and pharmacy<br>• Parks<br>• Transit stations |

**Table 2 – Mapping of Google mobility data to contact locations** (as defined by Prem et al.)

##### 3.7 Microdistancing

Interventions other than those that prevent people coming into contact with one another are thought to be important to COVID-19 transmission and epidemiology, such as maintaining interpersonal physical distance and the wearing of face coverings. We therefore implemented a "microdistancing" function to represent reductions in the rate of effective contact that is not attributable to persons visiting specific locations and so is not captured through Google mobility data. This microdistancing function reduces the values of all elements of the mixing matrices by a certain proportion. These time-varying functions multiplicatively scale the location-specific contact rate modifiers $s(t)$, $w(t)$ and $l(t)$.

#### 4 Simulation of local NPI implementation during Victoria's second wave

##### 4.1 School closures

The effect of Victorian school closures is captured through the timeline presented in Table 3.

| Date of change | Policy change | Modification applied to school contacts contribution to mixing matrix, $s(t)$ |
|---|---|---|
| From model start | Remote learning | 0.1 |
| 26th May | 400,000 school students return to school | 0.393 |
| 9th June | Remaining 618,000 school students return to school | 1 |

| | | |
|---|---|---|
| 9th July | Remote learning for stage 3 restrictions | 0.1 |

**Table 3 – Timeline used to implement Victorian school closure policies.** The function is applied to both metropolitan and regional clusters.

##### 4.2 Macrodistancing in workplaces and other locations

The functions applied here are determined by the Google mobility data according to Table 2, as described above, but are applied separately for each cluster. Because Google mobility data pertains to local government areas (LGAs), whereas health service clusters may receive patients from across the state, it was necessary to map mobility data to clusters. Health service clusters' overall mobility values in each location were calculated using a weighted average of LGA mobility values according to the historical pattern of the origin of patients presenting to services within each cluster.

As a hypothetical example, if 50% of patients historically presenting to Barwon South West health cluster services come from the City of Geelong, the mobility data for the City of Geelong will contribute 50% of the Google mobility estimate of Barwon South West.

Historical patterns of patient presentations by health service cluster were provided by the Victorian Department of Health and Human Services (DHHS).

##### 4.3 Microdistancing approach

In this application to Victoria, the microdistancing function $m(t)$ is comprised of two components: physical distancing and face coverings. Both physical distancing and face coverings micro-distancing are applied to the three non-household locations, such that the microdistancing function for non-household locations is given by:

$$m(t) = d(t)^2 \times f(t)^2$$

The two interventions are assumed to be independent and so are multiplicative. As for the macrodistancing functions, the two functions of time are squared to represent their effects on both the infector and the infectee in any potentially infectious interaction.

##### 4.4 Physical distancing

The physical distancing function $d(t)$ is a transposed and translated hyperbolic tan function. The parameters of this function were estimated by using maximum a posteriori inference, with priors that penalised large shape parameters (to avoid extremely rapid transitions). The proportions of respondents answering "always" to YouGov surveys of Victorian residents asking "Thinking about the last 7 days, about how many people from your household have you come into physical contact with (within 2 meters / 6 feet)?" were used as input data. Resulting parameters were: shape, 0.262764; lower asymptote, 0.2803973; upper asymptote; 0.4421819; and inflection point, 15$^{\text{th}}$ July. The resulting function is presented in Figure 5.

##### 4.5 Face coverings

Two separate face coverings microdistancing functions are employed, one for metropolitan and one for regional health service clusters. These functions were fitted using the same methods as for physical distancing, using YouGov data on Victorian residents' survey responses to the question "Thinking about the last 7 days, have you worn a face mask outside your home (e.g. when on public transport, going to a supermarket, going to a main road)?". Estimated parameters were: shape, 0.5261693; lower asymptote, 0.130469; upper asymptote, 0.9143849; and inflection point, 23$^{\text{rd}}$ July (consistent with the policy change in metropolitan Melbourne). This was applied directly to metropolitan clusters and translated ten days later for regional clusters, where face coverings were mandated from the 2$^{\text{nd}}$ August. The resulting function is presented in Figure 6.

#### 5 Between cluster mixing

The preceding section describes the creation of heterogeneous mixing matrices by age for each of the nine health service clusters individually. These mixing matrices are then combined to create a single time-

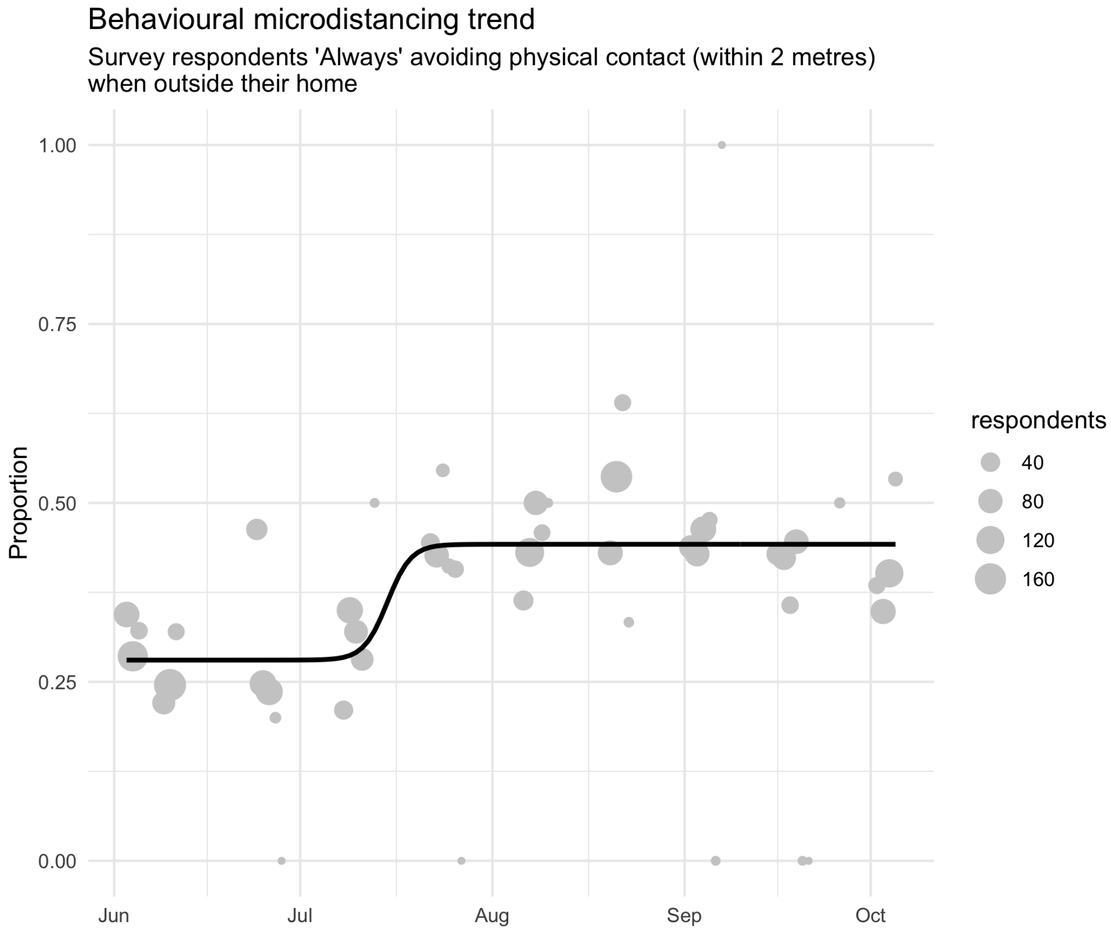

**Figure 5 – Physical distancing micro-distancing function (for all clusters) with data used for fitting.**

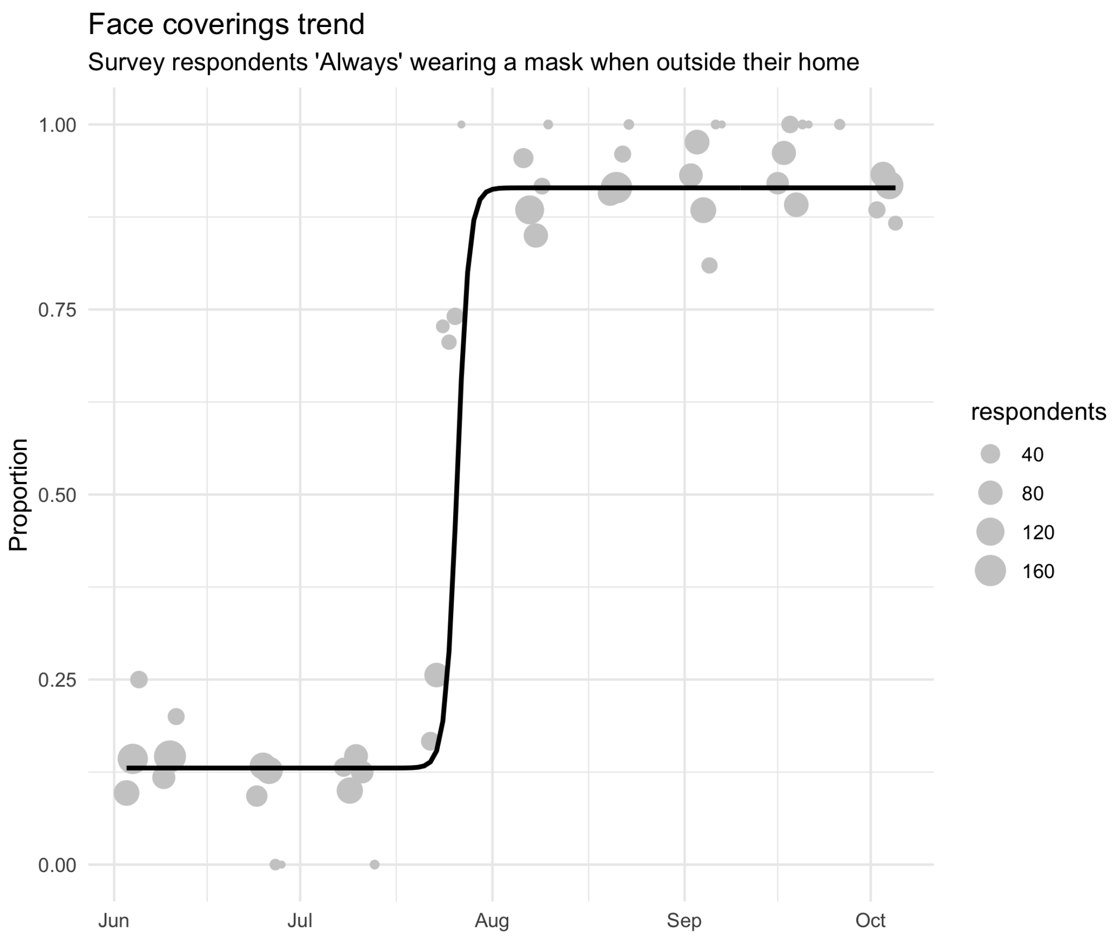

**Figure 6 – Face coverings micro-distancing function for metropolitan Melbourne clusters with data used for fitting.**

varying heterogeneous mixing matrix by cluster and age resulting in a 144 by 144 ($9 \times 16 = 144$) square mixing matrix. The force of infection for an index cluster is calculated from the mixing matrices of the age-assortative matrix for each of the clusters modelled. For clusters other than the index cluster, the mixing matrices are multiplied by a parameter that represents the extent of inter-cluster mixing. This is then added to the mixing matrix for the index cluster multiplied by $1 - 8 \times interclustermixing$ (because there are eight clusters other than the index cluster contributing to the mixing matrix) to create the final inter-cluster mixing matrix. The upper limit of prior of the *intercluster mixing* parameter is set to be considerably less than one ninth, to ensure most of the force of infection is contributed from the index cluster (Table 6).

### 6 Model initialisation

The model was commenced from approximately one to two weeks earlier than the actual beginning of Victoria's second wave (as determined by genomic analysis), in order that the distribution of infectious persons distributes naturally across compartments as the model approaches the actual beginning of Victoria's second wave in early June. The actual start date selected is the 14$^{\text{th}}$ May. The infectious seed needed at this time is then calibrated to ensure dynamics are realistic at the beginning of the second wave (see Table 6). The infectious seed is distributed evenly across metropolitan clusters, consistent with the epidemic's emergence from metrpolitan Melbourne.

### 7 Parameters

#### 7.1 Non-age-stratified parameters

| Parameter | Value | Rationale |
|---|---|---|
| Incubation period | Calibration parameter, truncated normal distribution, mean 5.5 days | Estimates of the incubation period have included 5.1 days, 5.2 days and 4.8 days [6] [7] [8] [9]. A systematic review [2] found that data are best fitted by a log-normal distribution (mean 5.8 days, CI 5.0 to 6.7, median 5.1 days). Our systematic review [10] found that estimates of the mean incubation period have varied from 3.6 to 7.4 days. |

Continuation of Table 4

| **Parameter** | **Value** | **Rationale** |
|---|---|---|
| Proportion of incubation period infectious | 50% | Infectiousness is considered to be present throughout a considerable proportion of the incubation period, based on analyses of confirmed source-secondary pairs [11] and early findings that the incubation period was similar to the serial interval [6]. The study of source-secondary pairs was also the primary reference cited by a review of the infectious period that identified studies that quantified the pre-symptomatic period, which concluded that the median pre-symptomatic period could range from less than one to four days [12]. |
| Active period (regardless of detection/isolation, for clinical strata 1 to 3) | Calibration parameter, truncated normal distribution, mean 6.5 days | This quantity is difficult to estimate, given that identified cases are typically quarantined. Studies in settings of high case ascertainment and an effective public health response have suggested a duration of greater than 5.5 days [9]. PCR positivity, which may continue for up to two to three weeks from the point of symptom onset [11] [12], is difficult to interpret and does not necessarily indicate infectiousness. Consistent with these findings, the duration infectious for asymptomatic persons has been estimated at 6.5 to 9.5 days [12] (although in our model, this would include the pre-symptomatic infectious period). |
| Proportion of infectious period before isolation or hospitalisation can occur | 0.333 | Assumed |
| Disease duration prior to admission for hospitalised patients not critically unwell (i.e. early active sojourn time, stratum 4) | 7.7 days | Mean value from ISARIC cohort, as reported on 4$^{th}$ October 2020 in Table 6 [13], and similar to the expected mean from earlier reports from ISARIC [14]. This cohort represents high-income countries better than low and middle-income countries, with the United Kingdom contributing data on the greatest number of patients, followed by France. Earlier estimates of this quantity from China included 4.4 days [6]. |
| Duration of hospitalisation if not critically unwell (late active sojourn time, stratum 4) | 11.6 days | Obtained from the Victorian Agency for Health Information |

Continuation of Table 4

| Parameter | Value | Rationale |
| --- | --- | --- |
| ICU duration (late active sojourn time, stratum 5) | 7.4 days | Obtained from the Victorian Agency for Health Information |
| Duration of time prior to ICU for patients admitted to ICU | 10.5 days | Calculated as the sum of the time from symptom onset to hospital admission (7.7 days above) plus the duration from hospital admission to ICU admission reported by October ISARIC report (2.8 days) [13]. |
| Relative infectiousness of asymptomatic persons (per unit time with active disease) | Calibration parameter, uniform distribution, range 0.15 to 0.7 | Assumed |
| Relative infectiousness of persons admitted to hospital or ICU | 0.2 | Assumed |
| Relative infectiousness of identified persons in isolation | 0.2 | Assumed |
| Proportion of hospitalised patients ever admitted to ICU | 0.16 | DHHS |

**Table 4 – Universal (non-age-stratified) model parameters.** Point estimates are used as model parameters except where ranges are indicated in calibration parameter table below.

##### *7.2 Age-specific parameters*

| Age group (years) | Clinical fraction[a] | Relative susceptibility to infection | Infection fatality rate | Proportion of symptomatic patients hospitalised |
|---|---|---|---|---|
| 0 to 4 | 0.29 | 0.36 | $3 \times 10^{-5}$ | 0.0777 |
| 5 to 9 | 0.29 | 0.36 | $1 \times 10^{-5}$ | 0.0069 |
| 10 to 14 | 0.21 | 0.36 | $1 \times 10^{-5}$ | 0.0034 |
| 15 to 19 | 0.21 | 1 | $3 \times 10^{-5}$ | 0.0051 |
| 20 to 24 | 0.27 | 1 | $6 \times 10^{-5}$ | 0.0068 |
| 25 to 29 | 0.27 | 1 | $1.3 \times 10^{-4}$ | 0.0080 |
| 30 to 34 | 0.33 | 1 | $2.4 \times 10^{-4}$ | 0.0124 |
| 35 to 39 | 0.33 | 1 | $4.0 \times 10^{-4}$ | 0.0129 |
| 40 to 44 | 0.40 | 1 | $7.5 \times 10^{-4}$ | 0.0190 |
| 45 to 49 | 0.40 | 1 | $1.21 \times 10^{-3}$ | 0.0331 |
| 50 to 54 | 0.49 | 1 | $2.07 \times 10^{-3}$ | 0.0383 |
| 55 to 59 | 0.49 | 1 | $3.23 \times 10^{-3}$ | 0.0579 |
| 60 to 64 | 0.63 | 1 | $4.56 \times 10^{-3}$ | 0.0617 |
| 65 to 69 | 0.63 | 1.41 | $1.075 \times 10^{-2}$ | 0.1030 |
| 70 to 74 | 0.69 | 1.41 | $1.674 \times 10^{-2}$ | 0.1072 |
| 75 and above | 0.69 | 1.41 | $5.748 \times 10^{-2}$, [b] | 0.0703 |
| Source/ rationale | Model fitting to age-distribution of early cases in China, Italy, Japan, Singapore, South Korea and Canada taken from upper-left panel of Figure 2b of [15]. | Conversion of odds ratios presented in Table S15 of Zhang et al. 2020 to relative risks using data presented in Table S14 of the same study [16].[c] | Estimated from pooled analysis of data from 45 countries from Table S3 of O'Driscoll et al [4]. Values consistent with previous estimates using serosurveys performed in Spain [17]. | Estimates from the Netherlands as the first wave of infections declined from 4th May to 21st July [18]. |

**Table 5 – Age-stratified parameter values.** Age-stratified parameters not varied during calibration, or varied through a common multiplier.

[a] Proportion of incident cases developing symptoms.

[b] Weighted average of IFR estimates for 70 to 79 and 80 and above age groups.

[c] Note the relative magnitude of these values are similar to those estimated by the analysis we use to estimate the age-specific clinical fraction.[15]

### 8 Calculation of outputs

#### 8.1 Incidence

Incidence is calculated as any transitions into the early active compartment (*"I"*).

#### 8.2 Hospital occupancy

This is calculated as the sum of three quantities:

1. All persons in the late active compartment in clinical stratum 4, representing those admitted to hospital but never critically unwell.
2. All persons in the late active compartment in clinical stratum 5, representing those currently admitted to ICU.
3. A proportion of the early active compartment in clinical stratum 5, representing those who will be admitted to ICU at a time in the future. This proportion is calculated as the quotient of *1)* the difference between the pre-ICU period and the pre-hospital period for clinical stratum 4, divided by *2)* the total pre-ICU period. That is, a proportion of the pre-ICU period is considered to represent patients in hospital who have not yet been admitted to ICU.

#### 8.3 ICU occupancy

This is calculated as all persons in the late active compartment in clinical stratum 4.

#### 8.4 Seropositive proportion

This is calculated as the proportion of the population in the recovered (*"R"*) compartment. Although very similar numerically to the attack rate, persons who died of COVID-19 are not included in the denominator.

#### 8.5 COVID-19-related mortality

This is calculated as all transitions representing death, exiting the model. This is implemented as depletion of the late active compartment.

##### 8.6 Notifications

Local case notifications are calculated as transitions from the early to the late active compartment for clinical strata 3 to 5.

#### 9 Calibration

We calibrated the model using the adaptive Metropolis algorithm described by Haario et al. [19]. A standard Metropolis algorithm with fixed proposal distribution parameters was used for the first 500 iterations to initiate the covariance matrix before the adaptive algorithm commenced.

##### 9.1 Rationale for cluster-specific targets

For all clusters (both metropolitan and regional), we included the time series of daily notifications for that cluster as a calibration target, using a normal distribution for the likelihood function. A normal distribution is preferred because the mapping process for the notifications for each cluster results in these quantities not being integer-valued.

In addition, we include time series for the following quantities at the state level. Because these quantities are counts, Poisson distributions are used in likelihood calculations:

- Daily new COVID-19 notifications
- Daily new hospital admissions
- Daily new ICU admissions
- Daily deaths

##### 9.2 Assigning targets to clusters

Hospital admissions and ICU admissions can be mapped directly to a health service cluster. Health service clusters include all health care (including public hospitals, private, rehab, acute, mental health, etc.) and some metropolitan services have changed cluster assignment over the years. Mapping was performed as at August 2020. However, for the other two indicators used (notifications and deaths), mapping was not possible because these events do not necessarily occur within a health service cluster. Therefore, the local government area (LGA) of residence of the person notified or dying is considered. Each notification and death is split proportionately across the health service clusters to which they would typically present, according to historical data on hospital presentations for each LGA provided by DHHS. (Note that only notifications are considered as calibration targets, although these considerations are relevant to the comparison between data and modelled outputs undertaken for validation purposes.)

| **Parameter name** | **Distribution type** | **Distribution parameters** |
|---|---|---|
| Incubation period (see Table 4) | Truncated normal | Mean 5.5 days, standard deviation 0.97 days, truncation $<$1 day |
| Infectious period (for clinical strata 1 to 3) (see Table 4) | Truncated normal | Mean 6.5 days, standard deviation 0.77 days, truncation $<$1 day |
| Risk of infection per contact (before adjustments) | Uniform | 0.015 to 0.06 |
| Intercluster mixing (proportion of infection contributed by each non-index cluster) | Uniform | Range 0.005 to 0.05 |
| Infectious seed | Uniform | 10 to 30 |
| Seasonal forcing (relative change to contact probability from mid-summer to mid-winter) | Uniform | Range 0 to 0.5 |

Continuation of Table 6

| **Parameter name** | **Distribution type** | **Distribution parameters** |
|---|---|---|
| Clinical fraction adjuster | Truncated normal | Mean 1, standard deviation 0.2, truncation $<$0.5 |
| Relative infectiousness of asymptomatic patients | Uniform | Range 0.15 to 0.7 |
| Hospitalisation proportions adjuster | Normal | Range 0.5 to 3 |
| Infection fatality rate adjuster | Normal | Range 0.5 to 4 |
| Proportion of symptomatic cases that would be detected with daily per capita testing rate of one per thousand | Uniform | Range 0.2 to 0.5 |
| Disease duration prior to admission to ICU (early disease, stratum 5 sojourn time) | Truncated normal | Mean 12.7 days, standard deviation 4 days, truncated $<$3 days |
| Effect of physical distancing | Normal | Range 0 to 0.5 |
| Effect of face coverings | Normal | Range 0 to 0.5 |

Continuation of Table 6

| Parameter name | Distribution type | Distribution parameters |
|---|---|---|
| Five parameters to adjust the probability of infection given contact in each of the four metropolitan clusters for Barwon South West | Truncated normal | Mean 1, standard deviation 0.5, truncation $<$0.5 |
| One parameter to adjust the probability of infection given contact in all of the remaining four regional clusters | Truncated normal | Mean 1, standard deviation 0.5, truncation $<$0.5 |

**Table 6 – Calibration parameters.**

##### 9.3 Variation of age-specific proportion parameters using "adjuster" parameters

The following sections describe age-specific parameters that were varied during calibration. These proportion parameters are modified through "adjuster" parameters that are not strictly multipliers, but are rather implemented in such a way as to scale the base parameter value while ensuring that the adjusted parameter remains a proportion (with range zero to one). In each of these cases, the adjuster parameters can be considered as multiplicative factors that are applied to the odds ratio that is equivalent to the baseline proportion to be adjusted. Specifically, the adjusted proportion is equal to:

$$\frac{proportion \times adjuster}{proportion \times (adjuster - 1) + 1}$$

##### 9.4 Variation of the proportion of patients symptomatic

The modelled proportion of patients symptomatic differs by age group. However, given that this quantity remains highly uncertain and may vary between settings, it is varied during calibration. A single adjuster is used to increase or decrease each value for each age group.

##### 9.5 Variation of the proportion of patients hospitalised

The modelled proportion of patients hospitalised similarly differs by age group, and is also likely to vary between settings. A single adjuster is used to increase or decrease each value for each age group.

##### 9.6 Variation of infection fatality rate

The infection fatality rate (risk of death given infection) is considered a more stable quantity than the case fatality rate. However, it is still likely to vary considerably between settings and so is included as a calibration parameter which adjusts each age-specific IFR by the same value. Because the epidemic in Victoria has been characterised by high rates of transmission and disease in aged care, at baseline we assign a prior centred at a value greater than one.

#### 10 Likelihood function

Likelihood functions are derived from comparing model outputs to target data at each time point nominated for calibration.

The composite likelihood function is given formally as:

$$\prod_{t} n_t(\theta) d_t(\theta) h_t(\theta) i_t(\theta) \times \prod_{t,g} n_{t,g}(\theta, \sigma)$$

where $_t$ indexes the date, $_g$ indexes the cluster, $n_t$ refers to daily new notifications, $d_t$ to daily deaths, $h_t$ to daily new hospitalisations and $i_t$ to daily new ICU admissions. Each state-wide component uses a Poisson distribution (e.g. $n_t(\theta) = Poiss(\nu_t(\theta))$, where $\nu_t(\theta)$ is the number of notifications simulated by the model at date t under parameter set $\theta$), whereas each $n_{t,g}$ uses a normal likelihood distribution (because these targets are not integer-valued). $\sigma$ is ratio of the peak of each cluster-specific notification to the corresponding standard deviation of each of the normal distributions used in calculating their contribution to the likelihood. This was included as a calibration parameter to improve calibration efficiency.

#### 11 Ordinary differential equations

For the clearest description of the model, we refer the reader to our code repository, because our object-oriented approach to software development is intended to be highly transparent and readable. For those who prefer dynamical systems such as this presented in the form of ordinary differential equations, we present the following.

$$\frac{dS_{a,g}}{dt} = -\lambda_{a,g}(t) \times \sigma_a \times S_{a,g}$$
$$\frac{dE_{a,g}}{dt} = \lambda_{a,g}(t) \times \sigma_a \times S_{a,g} - \alpha E_{a,g}$$
$$\frac{dP_{a,c,g}}{dt} = p_{a,c}(t) \times \alpha E_{a,g} - \nu P_{a,c,g}$$
$$\frac{dI_{a,c,g}}{dt} = \nu P_{a,c,g} - \gamma_c I_{a,c,g}$$
$$\frac{dL_{a,c,g}}{dt} = \gamma_c I_{a,c,g} - \delta_{a,c} L_{a,c,g} - \mu_{a,c} L_{a,c,g}$$
$$\frac{dR_{a,g}}{dt} = \sum_c \delta_{a,c} L_{a,c,g}$$

where

$$\lambda_{a,g} = \beta(t) \sum_{g'} \mathbf{G}_{g,g'} \sum_{j,c} \frac{\varepsilon P_{j,c,g'}(t) + \iota_c I_{j,c,g'}(t) + \kappa_c L_{j,c,g'}(t)}{N_{j,g'}(t)} C_{a,j}(t)$$

$$\sum_c p_{a,c}(t) = 1, \forall t \in \mathbb{R}$$

$$\mathbf{C}_0 = \mathbf{C}_H + \mathbf{C}_S + \mathbf{C}_W + \mathbf{C}_L$$

$$\mathbf{C}_g(t) = \mathbf{C}_H + s_g(t)^2 \mathbf{C}_S + w_g(t)^2 \mathbf{C}_W + l_g(t)^2 \mathbf{C}_L$$

$$l_g(t) = \frac{re_g(t) + gr_g(t) + pa_g(t) + tr_g(t)}{4}$$

| Symbol | Explanation |
|---|---|
| $S$ | Persons susceptible to infection |
| $E$ | Persons in the non-infectious incubation period |
| $P$ | Persons in the incubation period |
| $I$ | Persons in the early active disease period, before isolation or hospitalisation may occur |
| $L$ | Persons in the late active disease period, after isolation or hospitalisation may have occurred |
| $R$ | Persons in the recovered period, from which re-infection cannot occur |

| Symbol | Explanation |
|---|---|
| $t$ | Time |
| $a$ | Compartment of age group a |
| $c$ | Compartment of clinical stratification c |
| $g$ | Compartment of geographical cluster stratificiation g |
| $\alpha$ | Rate of progression from non-infectious to infectious incubation period |
| $\nu$ | Rate of progression from infectious incubation to early active disease |
| $\gamma$ | Rate of progression from early active disease to late active disease |
| $\mu$ | Rate of disease-related death |
| $\varepsilon$ | Relative infectiousness of pre-symptomatic compartment |
| $\iota$ | Clinical stratification infectiousness vector for early active compartment |
| $\kappa$ | Clinical stratification infectiousness vector for late active compartments |
| $\beta(t)$ | Seasonally adjusted probability of infection per contact between an infectious and susceptible individual |
| $j$ | Infectious populations |
| $p$ | Proportion progressing to each clinical stratification |
| **G** | Square matrix of dimensions $9 \times 9$ (for nine clusters) with values of mixing parameter for the off-diagonal elements and values of $1 - 8 \times mixingparameter$ |

| **Symbol** | **Explanation** |
|---|---|
| $\mathbf{C}$ | Mixing matrix |
| $\mathbf{H}$ | Household contribution to mixing matrix |
| $\mathbf{W}$ | Workplace contribution to mixing matrix |
| $\mathbf{O}$ | Other locations contribution to mixing matrix |
| $\mathbf{S}$ | Schools contribution to mixing matrix |
| $l$ | Other locations macrodistancing function of time |
| $w$ | Function fit to Google mobility data for workplaces |
| $s$ | Function fit to Google mobility data for schools |
| $re$ | Function fit to Google mobility data for retail and recreation |
| $gr$ | Function fit to Google mobility data for grocery and pharmacy |
| $pa$ | Function fit to Google mobility data for parks |
| $tr$ | Function fit to Google mobility data for transit stations |

#### 12 Supplemental Output Figures

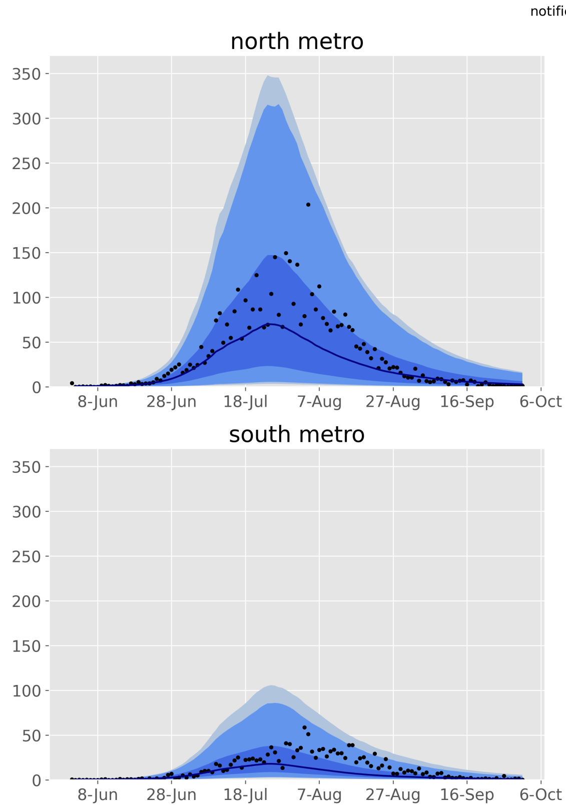

**Figure 7 – Calibration fit to daily time series of notifications for each metropolitan health service cluster.** Daily confirmed cases (black dots) overlaid on the median modeled detected cases (dark blue line), with shaded areas representing the 25$^{th}$ to 75$^{th}$ centile (mid blue), 2.5$^{th}$ to 97.5$^{th}$ centile (light blue) and 1$^{st}$ to 99$^{th}$ centile (faintest blue) of estimated detected cases.

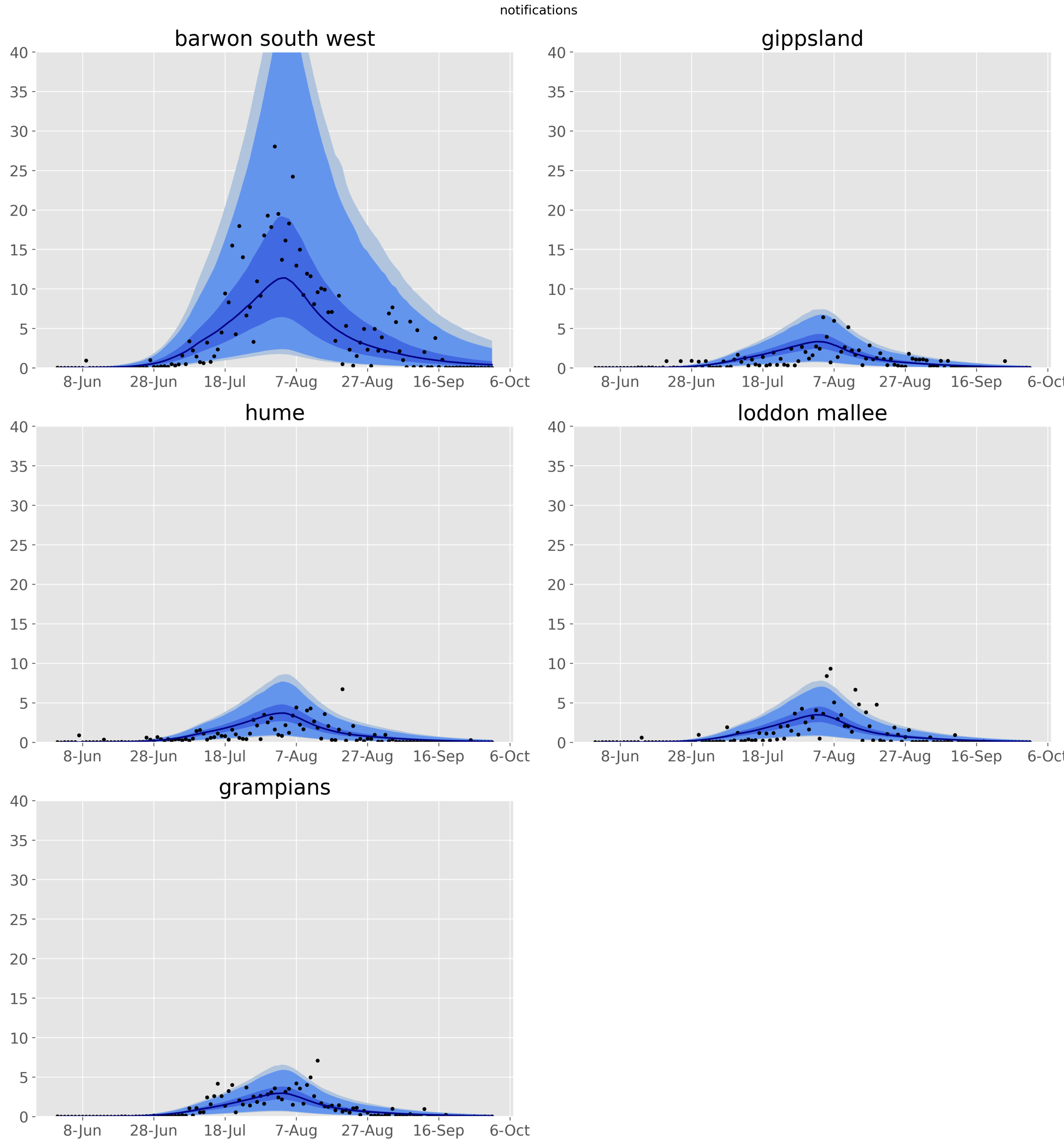

**Figure 8 – Calibration fit to daily time series of notifications for each regional health service cluster.** Daily confirmed cases (black dots) overlaid on the median modeled detected cases (dark blue line), with shaded areas representing the $25^{th}$ to $75^{th}$ centile (mid blue), $2.5^{th}$ to $97.5^{th}$ centile (light blue) and $1^{st}$ to $99^{th}$ centile (faintest blue) of estimated detected cases.

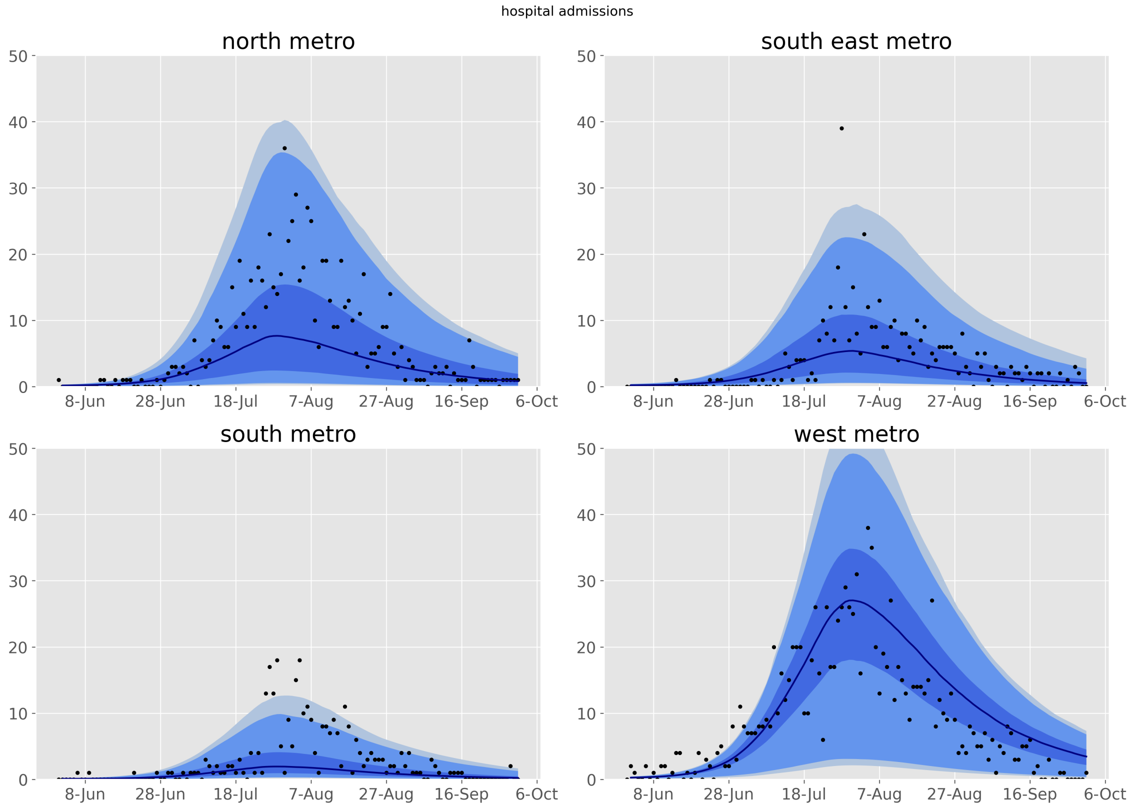

**Figure 9 – Validation fit to daily time series of hospitalisations for each metropolitan health service cluster.** Daily confirmed cases (black dots) overlaid on the median modeled detected cases (dark blue line), with shaded areas representing the $25^{th}$ to $75^{th}$ centile (mid blue), $2.5^{th}$ to $97.5^{th}$ centile (light blue) and $1^{st}$ to $99^{th}$ centile (faintest blue) of estimated detected cases.

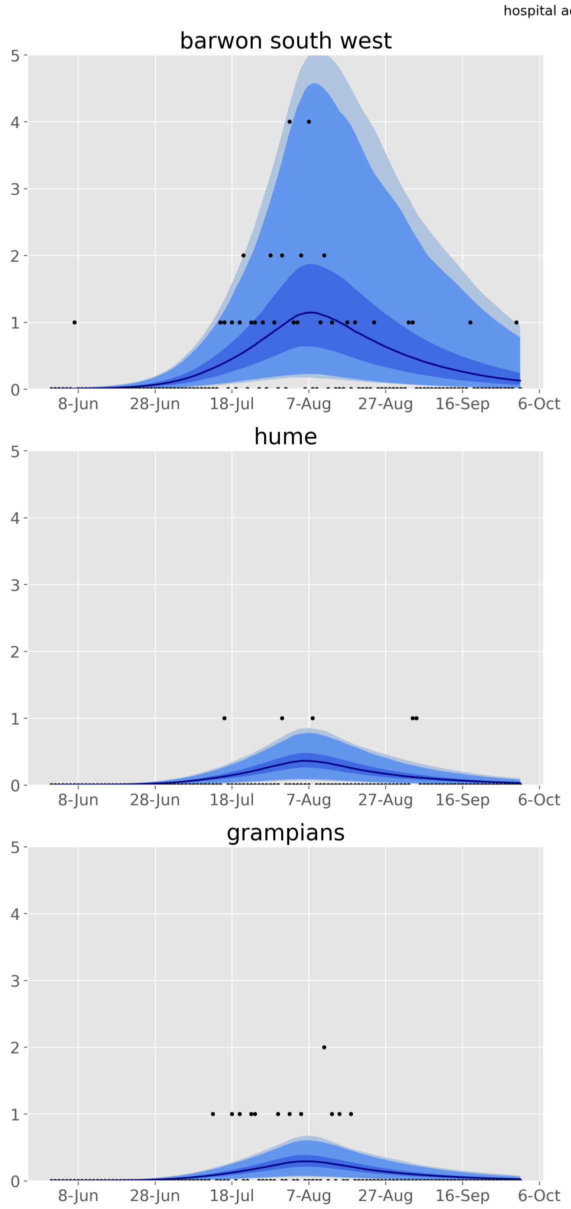

**Figure 10 – Validation fit to daily time series of hospitalisations for each regional health service cluster.** Daily confirmed cases (black dots) overlaid on the median modeled detected cases (dark blue line), with shaded areas representing the 25$^{th}$ to 75$^{th}$ centile (mid blue), 2.5$^{th}$ to 97.5$^{th}$ centile (light blue) and 1$^{st}$ to 99$^{th}$ centile (faintest blue) of estimated detected cases.

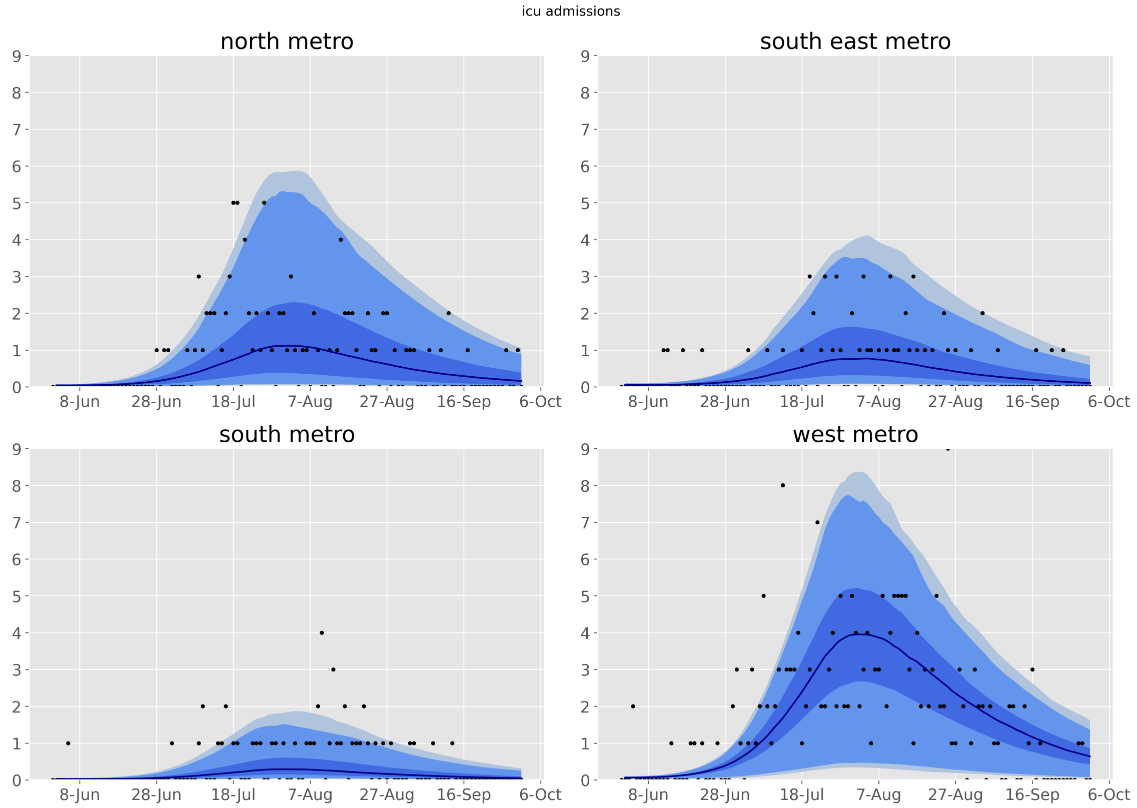

**Figure 11 – Validation fit to daily time series of ICU admissions for each metropolitan health service cluster.** Daily confirmed cases (black dots) overlaid on the median modelled detected cases (dark blue line), with shaded areas representing the $25^{th}$ to $75^{th}$ centile (mid blue), $2.5^{th}$ to $97.5^{th}$ centile (light blue) and $1^{st}$ to $99^{th}$ centile (faintest blue) of estimated detected cases.

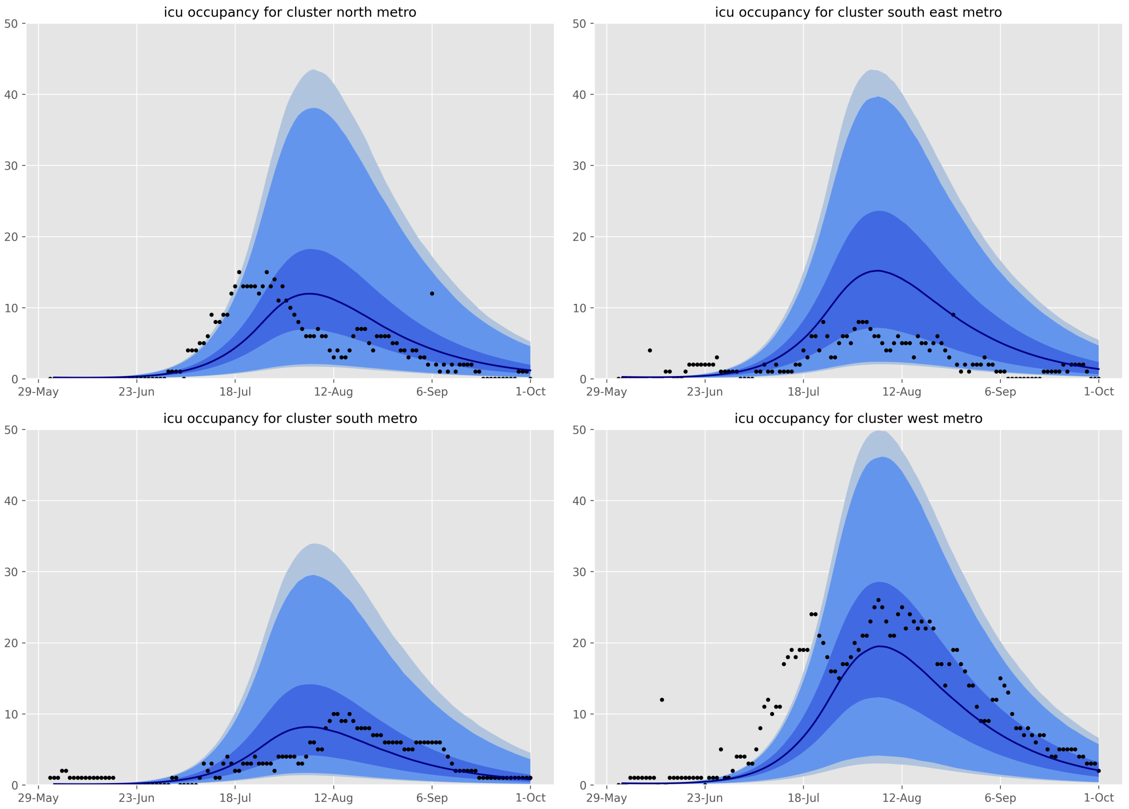

**Figure 12 – Validation fit to ICU occupancy for each metropolitan health service cluster.** Daily occupancy values (black dots) overlaid on the median modelled detected cases (dark blue line), with shaded areas representing the $25^{th}$ to $75^{th}$ centile (mid blue), $2.5^{th}$ to $97.5^{th}$ centile (light blue) and $1^{st}$ to $99^{th}$ centile (faintest blue) of estimated detected cases. Note that this data was a particularly poor validation/calibration target because of the number of inter-ICU transfers that affected the cluster-specific bed occupancy values.

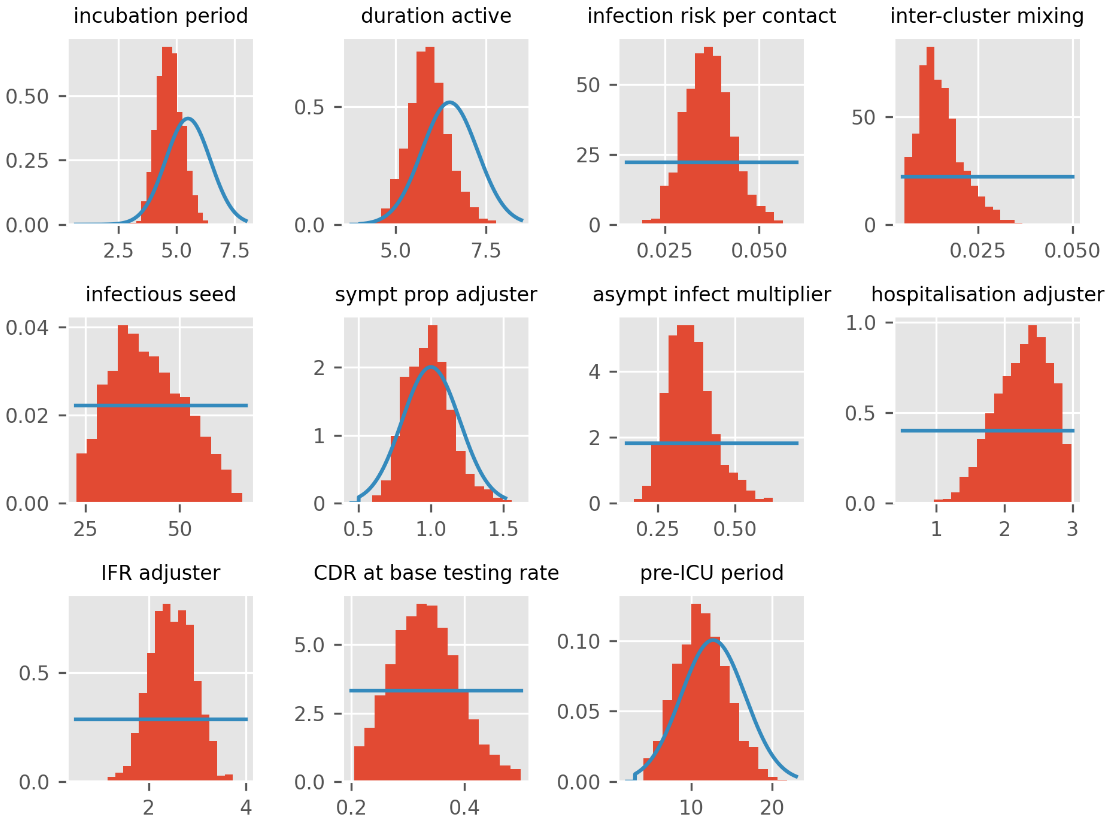

**Figure 13 – Histograms of state-wide epidemiological parameter posteriors, other than key parameters of interest** (presented in main manuscript).

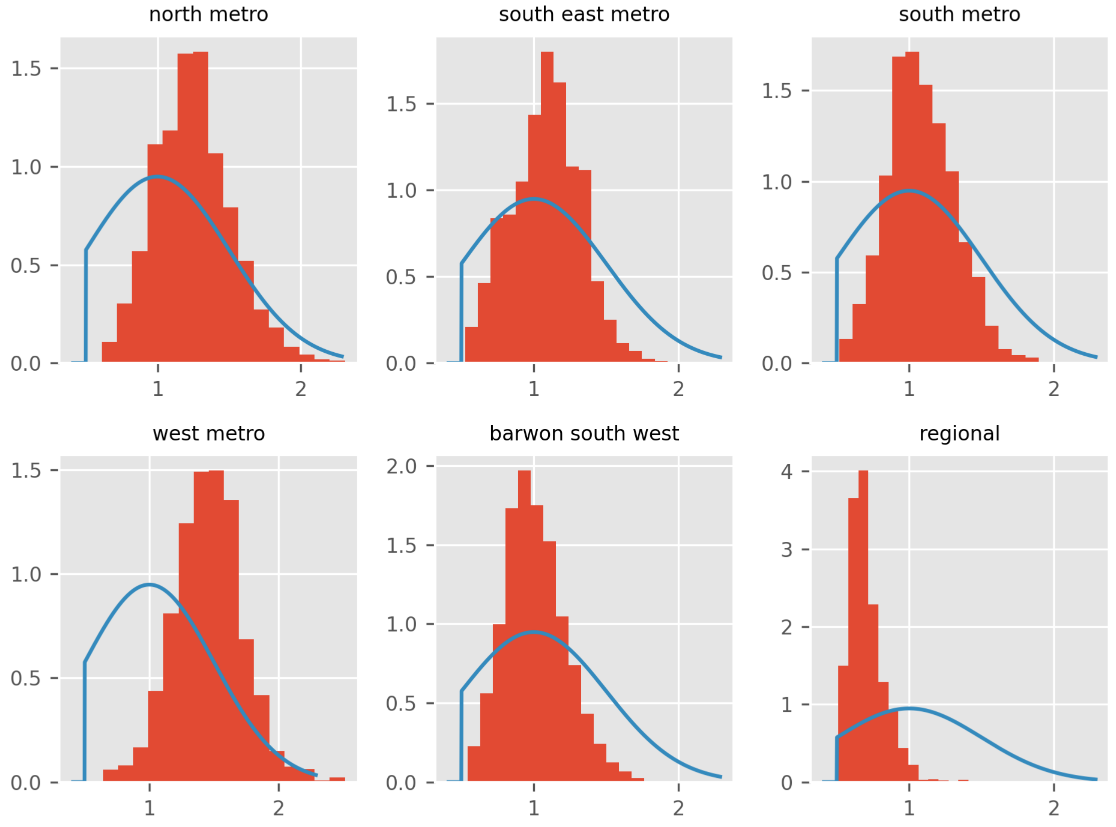

**Figure 14 – Histograms of cluster-specific contact rate modifier parameter posteriors.**

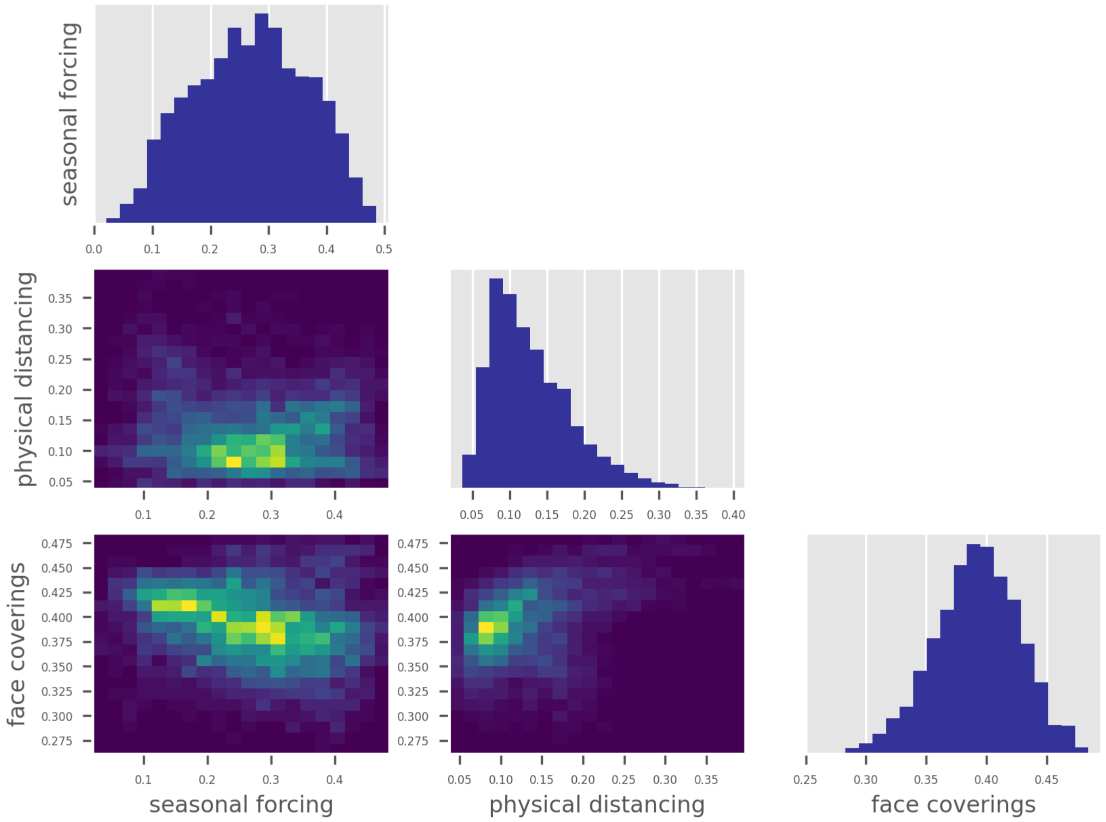

**Figure 15 – Correlation matrix for key epidemiological parameters of interest.**

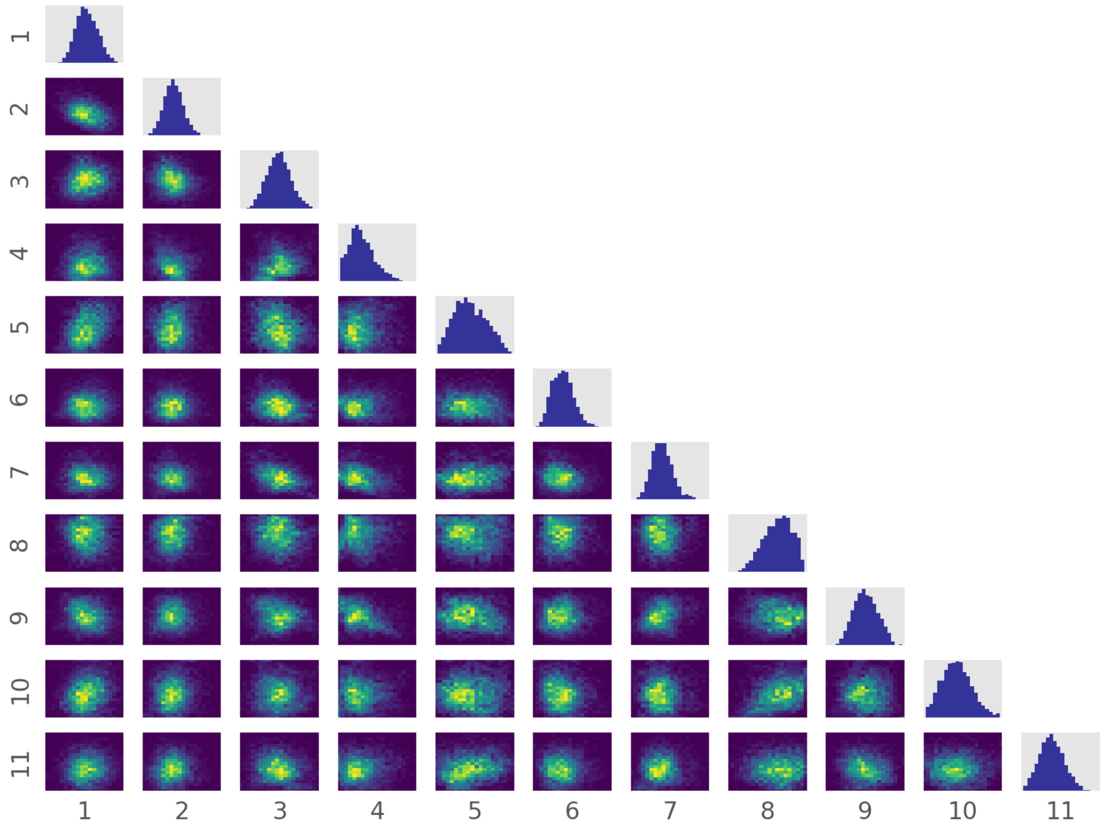

**Figure 16** – **Correlation matrix for other state-wide epidemiological parameters.** Parameters are: 1, incubation period; 2, duration active; 3, infection risk per contact; 4, inter-cluster mixing; 5, infectious seed; 6, sympt prop adjuster; 7, asympt infect multiplier; 8, hospitalisation adjuster; 9, IFR adjuster; 10, CDR at base testing rate; 11, pre-ICU period.

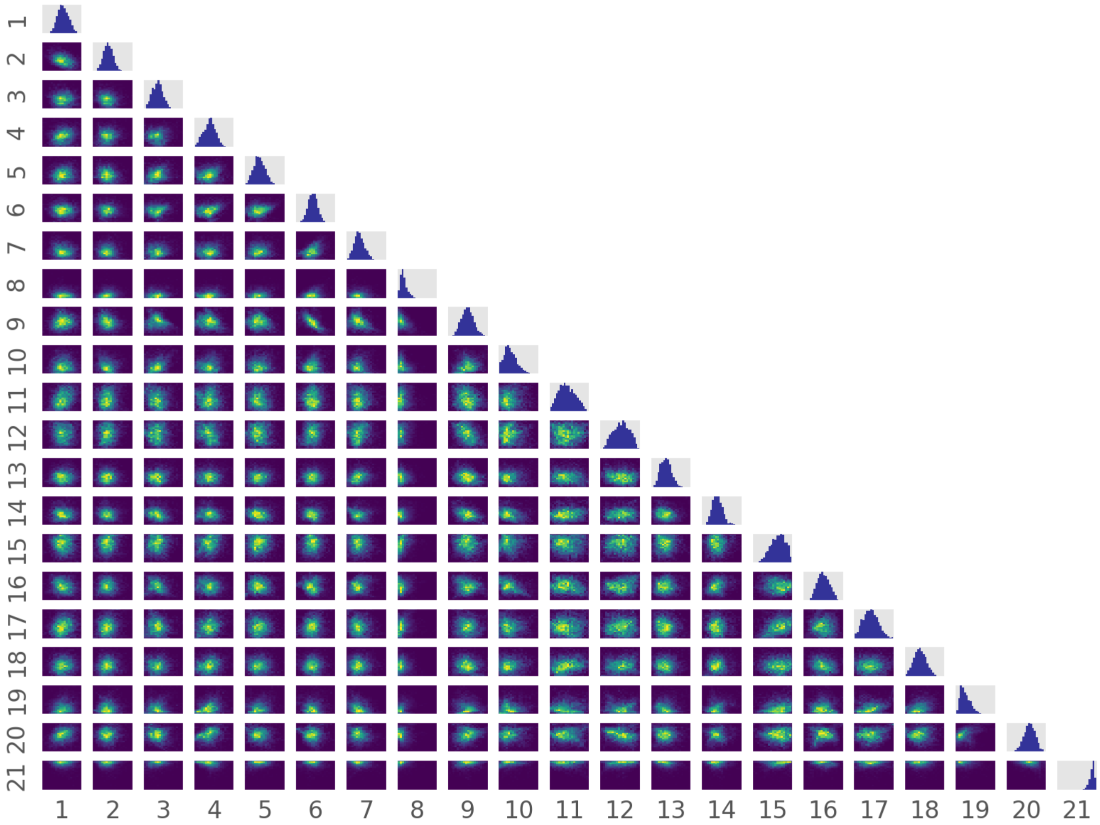

**Figure 17 – Correlation matrix for all parameters.** Parameters are: 1, incubation period; 2, duration active; 3, north metro; 4, south east metro; 5, south metro; 6, west metro; 7, barwon south west; 8, regional; 9, infection risk per contact; 10, inter-cluster mixing; 11, infectious seed; 12, seasonal forcing; 13, sympt prop adjuster; 14, asympt infect multiplier; 15, hospitalisation adjuster; 16, IFR adjuster; 17, CDR at base testing rate; 18, pre-ICU period; 19, physical distancing; 20, face coverings; 21, target output ratio.

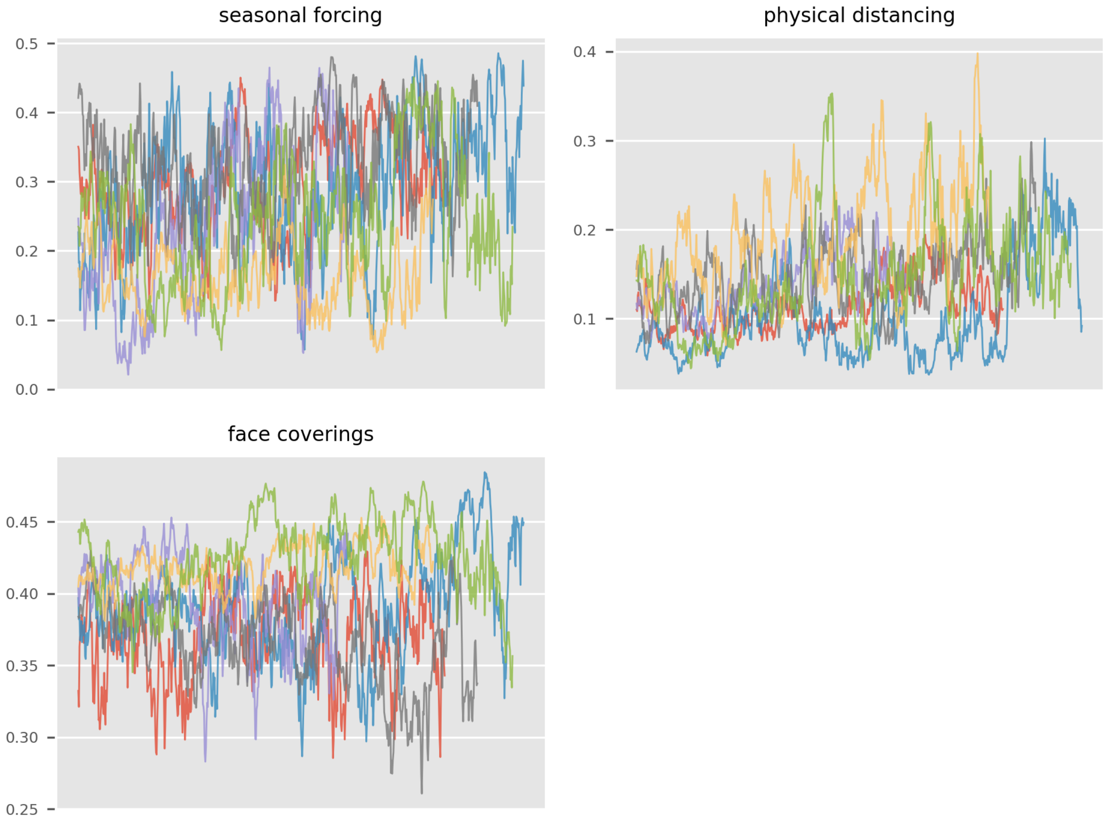

**Figure 18 – Parameter progression traces for key estimation parameters.**

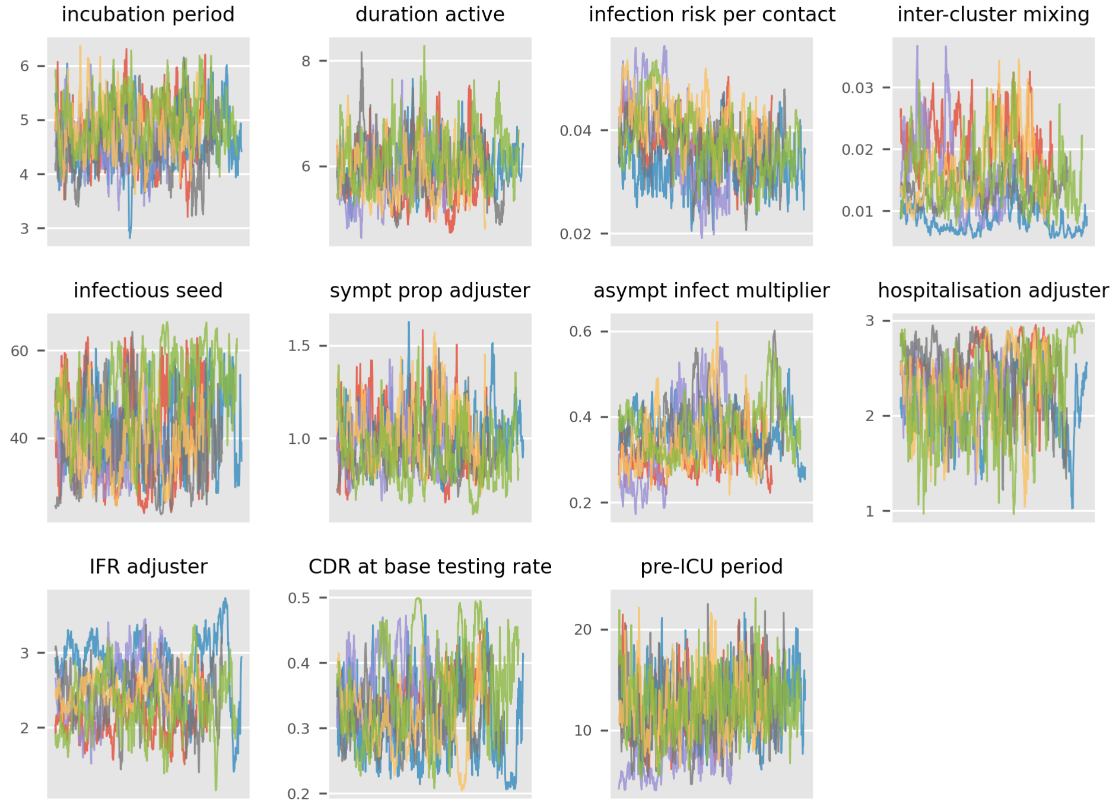

**Figure 19 – Parameter progression traces for epidemiological parameters.**

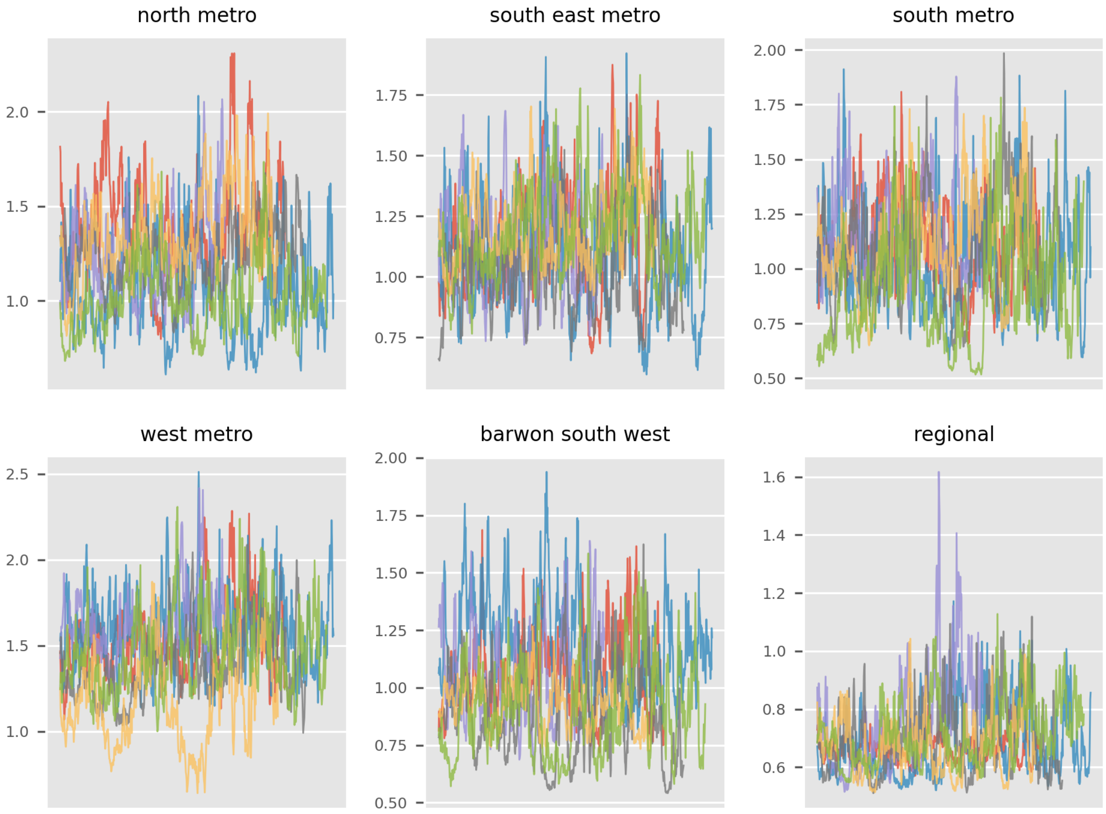

**Figure 20 – Parameter progression traces for cluster contact rate modifier parameters.**

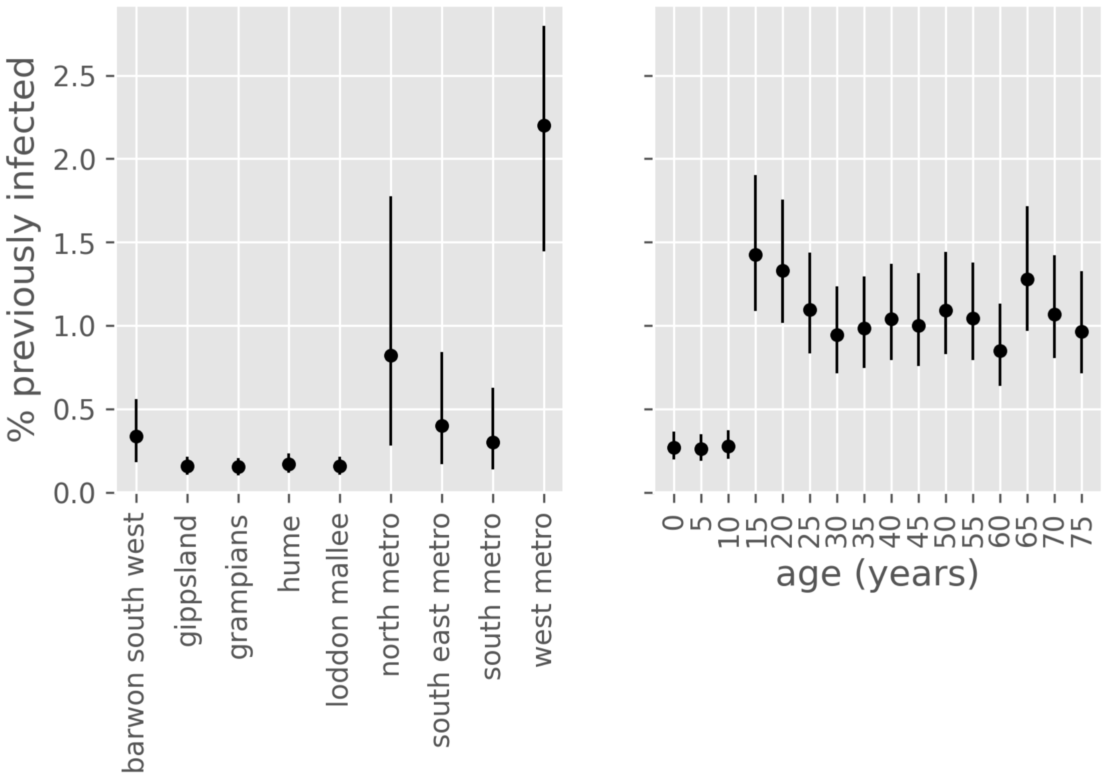

**Figure 21 – Estimated proportion of population recovered from COVID-19 at 1st October 2020, by age group and health service cluster.** Point estimates with associated 50% credible intervals. Values are negligibly different from attack rates, except that deaths are excluded from the denominator. Infections from first wave not considered.

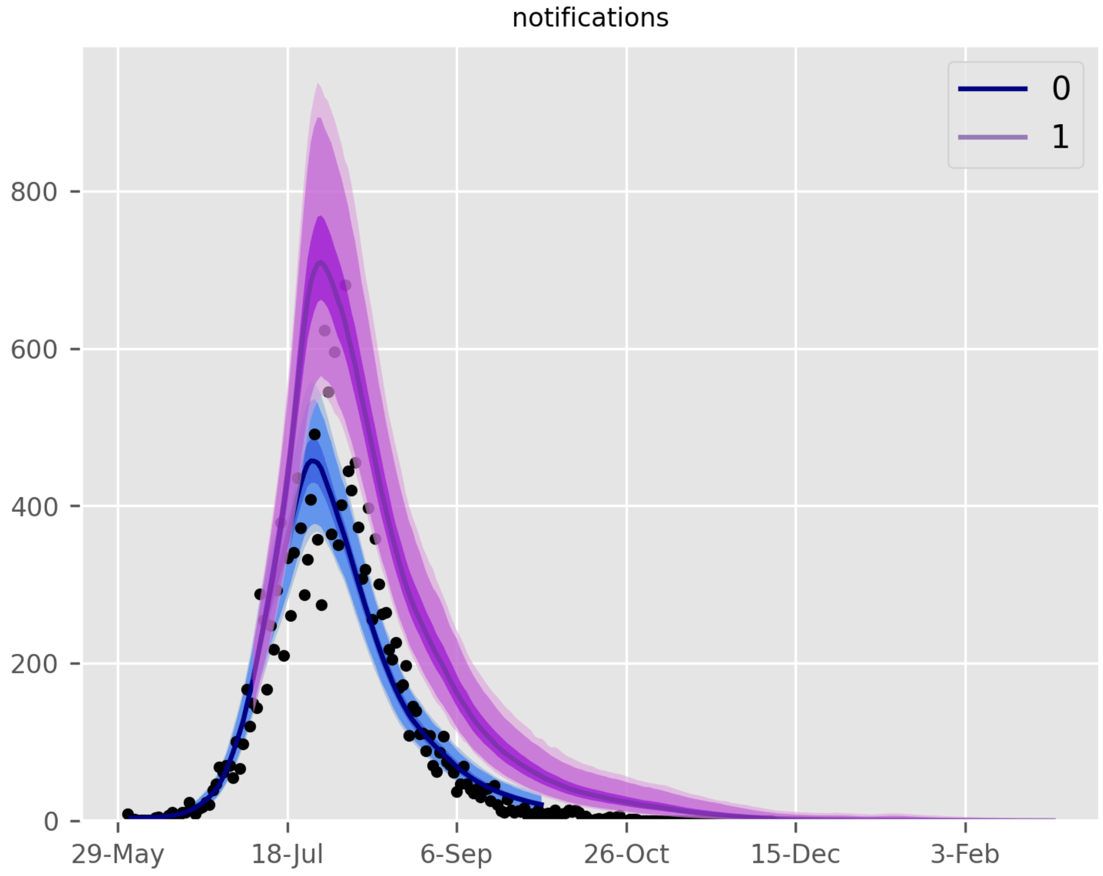

**Figure 22 – Scenario plot showing only baseline calibration and school re-opening scenario.** Scenarios are: blue, baseline; purple, schools re-opened from 7th July.
